# On Multiply Robust Mendelian Randomization (MR^2^) With Many Invalid Genetic Instruments

**DOI:** 10.1101/2021.10.21.21265317

**Authors:** Baoluo Sun, Zhonghua Liu, Eric Tchetgen Tchetgen

## Abstract

Mendelian randomization (MR) is a popular instrumental variable (IV) approach, in which genetic markers are used as IVs. In order to improve efficiency, multiple markers are routinely used in MR analyses, leading to concerns about bias due to possible violation of IV exclusion restriction of no direct effect of any IV on the outcome other than through the exposure in view. To address this concern, we introduce a new class of Multiply Robust MR (MR^2^) estimators that are guaranteed to remain consistent for the causal effect of interest provided that at least one genetic marker is a valid IV without necessarily knowing which IVs are invalid. We show that the proposed MR^2^ estimators are a special case of a more general class of estimators that remain consistent provided that a set of at least *k*^†^ out of *K* candidate instrumental variables are valid, for *k*^†^ *≤ K* set by the analyst ex ante, without necessarily knowing which IVs are invalid. We provide formal semiparametric theory supporting our results, and characterize the semiparametric efficiency bound for the exposure causal effect which cannot be improved upon by any regular estimator with our favorable robustness property. We conduct extensive simulation studies and apply our methods to a large-scale analysis of UK Biobank data, demonstrating the superior empirical performance of MR^2^ compared to competing MR methods.

## 1 Introduction

Mendelian randomization (MR) is an instrumental variable (IV) approach with growing popularity in epidemiology studies. In MR, one aims to establish a causal association between a given exposure and an outcome of interest in the presence of possible unmeasured confounding, by leveraging one or more genetic markers defining the IV (Davey Smith and Ebrahim, 2003; Lawlor et al., 2008). In order to be a valid IV, a genetic marker must satisfy the following key conditions:

a. It must be associated with the exposure.
b. It must be independent of any unmeasured confounder of the exposure-outcome relationship.
c. There must be no direct effect of the genetic marker on the outcome which is not fully mediated by the exposure in view.

Assumption (c) also known as the exclusion restriction is rarely credible in the MR context as it requires complete understanding of the biological mechanism by which each genetic marker influences the outcome. Such a priori knowledge may be unrealistic in practice due to the possible existence of unknown pleitropic effects of the markers (Little and Khoury, 2003; Davey Smith and Ebrahim, 2003; Lawlor et al., 2008). Violation of assumption (b) can also occur due to linkage disequilibrium or population stratification (Lawlor et al., 2008). Possible violation or near violation of assumption (a) known as the weak instrumental variable problem also poses an important challenge in MR as individual genetic effects on complex traits can be weak.

There has been tremendous interest in the development of methods to detect and account for violation of IV assumptions (a)-(c), primarily in multiple-IV settings under standard linear outcome and exposure models. Literature addressing violation of assumption (a) is arguably the most developed and extends to possible nonlinear models under a generalized methods of moments framework; notable papers of this rich literature include Staiger and Stock (1997); Stock and Wright (2000); Stock et al. (2002); Chao and Swanson (2005); Newey and Windmeijer (2009). A growing literature has likewise emerged on methods to address violations of assumptions (b) and (c), a representative sample of which includes Bowden et al. (2015, 2016); Kang et al. (2016); Windmeijer et al. (2019); Liu et al. (2020b); Tchetgen Tchetgen et al. (2021); Ye et al. (2021). The current paper is perhaps most closely related to the works of Kang et al. (2016); Guo et al. (2018); Windmeijer et al. (2019). Specifically, Kang et al. (2016) developed a penalized regression approach that can recover valid inferences about the causal effect of interest provided fewer than fifty percent of genetic markers are invalid IVs (known as majority rule); Windmeijer et al. (2019) improved on the penalized approach, including a proposal for standard error estimation lacking in Kang et al. (2016). In an alternative approach, Han (2008) established that the median of multiple estimators of the effect of exposure obtained using one instrument at the time is a consistent estimator under majority rule and that IVs cannot have direct effects on the outcome unless the IVs are uncorrelated. Guo et al. (2018) proposed two stage hard thresholding (TSHT) with voting, which is consistent for the causal effect under linear outcome and exposure models, and a plurality rule which can be considerably weaker than the majority rule. The plurality rule is defined in terms of regression parameters encoding (i) the association of each invalid IV with the outcome and that encoding (ii) the association of the corresponding IV with the exposure. The condition effectively requires that the number of valid IVs is greater than the largest number of invalid IVs with equal ratio of regression coefficients (i) and (ii). Furthermore, they provide a simple construction for 95% confidence intervals to obtain inferences about the exposure effect which are guaranteed to have correct coverage under the plurality rule. Importantly, in these works, a candidate IV may be invalid either because it violates the exclusion restriction, or because it shares an unmeasured common cause with the outcome, i.e. either (b) or (c) fails. Both the penalized approach and the median estimator may be inconsistent if majority rule fails, while TSHT may be inconsistent if plurality rule fails. It is important to note that because confidence intervals for the causal effect of the exposure obtained by Windmeijer et al. (2019) and Guo et al. (2018) rely on a consistent model selection procedure, such confidence intervals fail to be uniformly valid over the entire model space (Leeb and Pötscher, 2008; Guo et al., 2018).

Interestingly, Kang et al. (2016); Guo et al. (2018) and Windmeijer et al. (2019) effectively tackle the task of obtaining valid inferences about a confounded causal effect in the presence of invalid IVs as a model selection problem. Specifically, they aim to correctly identify which candidate IVs are invalid, while simultaneously obtaining valid MR inferences using selected valid IVs, arguably a more challenging task than may be required to obtain causal inferences that are robust to the presence of invalid IVs without necessarily knowing which IVs are invalid. In contrast, the methods proposed in this paper by-pass the model selection step altogether to deliver a regular and asymptotically linear multiply robust MR (MR^2^) estimator of the causal parameter of interest, provided at least one genetic instrument satisfies IV assumptions (a)-(c) without knowledge of the invalid IVs. Therefore, our estimators can be consistent with a single valid IV even when majority and plurality rules fail to hold. Furthermore, our estimators are regular and asymptotically linear, and therefore can be used to obtain uniformly valid 95% confidence intervals in large samples. We further show that the proposed estimators are a special case of a more general class of estimators that remain consistent provided that *k*^†^ out of *K* candidate instrumental variables are valid, for *k*^†^ *≤ K* set by the analyst ex ante, without necessarily knowing which *K −k*^†^ are invalid IVs. A necessary trade-off between more stringent IV relevance requirements in exchange for less stringent IV exclusion restriction requirements is revealed by increasing the value of *k*^†^, thus providing formal quantification of the efficiency cost one must incur in exchange for increased robustness. We characterize a semiparametric efficiency bound which cannot be improved upon by any regular and asymptotically linear estimator which shares the robustness property of our class of estimators.

The rest of the article proceeds as follows. In Section 2, we introduce notation and definitions. In Section 3, we describe our novel identification strategy with many independent invalid IVs. In Section 4, we perform extensive simulation studies to demonstrate the empirical performance of our proposed MR^2^ estimator along with competing ones, and apply our method in Section 5 to estimate the average causal effects of body mass index on systolic and diastolic blood pressure using large-scale UK Biobank data. In Sections 6 and 7, we discuss generalizations to the setting with correlated invalid IVs and partial identification respectively. We conclude with a brief discussion in Section 8. The R package for the proposed method is publicly available at: https://github.com/zhonghualiu/MRSquared.

## 2 Notation and Definitions

Suppose that one has observed *n* i.i.d. realizations of a vector (*Y, A, G*_1_, …, *G*_*K*_) where *A* is an exposure, 𝒢 = {*G*_*k*_ : *k* = 1, …*K*} are candidate genetic IVs and *Y* is the outcome. Let *U* denote an unmeasured confounder (possibly multivariate) of the effect of *A* on *Y*. 𝒢 *\G*_*k*_ denotes the set of candidate genetic IVs excluding *G*_*k*_. Then, *G*_*k*_ is said to be a valid instrumental variable provided it fulfills the following three conditions:

### Assumption 1.

IV relevance: *G*_*k*_ */ ⊥A*|𝒢 *\G*_*k*_;

### Assumption 2.

IV independence: *G*_*k*_*⊥U*|𝒢 *\G*_*k*_;

### Assumption 3.

Exclusion restriction: *G*_*k*_*⊥Y* |*A, U*, 𝒢 *\G*_*k*_.

The first condition ensures that the IV is a correlate of the exposure. The second condition states that the IV is independent of all unmeasured confounders of the exposure-outcome association, while the third condition formalizes the assumption of no direct effect of *G*_*k*_ on *Y* not mediated by *A* (presuming Assumption 2 holds). The causal diagram in Figure 1 encodes these three assumptions and therefore provides a graphical representation of the IV model. It is well known that while a valid IV satisfying assumptions 1-3, i.e. the causal diagram in Figure 1, suffices to obtain a valid statistical test of the sharp null hypothesis of no individual causal effect, the population average causal effect is itself not point identified with a valid IV without an additional assumption. Consider the following condition:

**Figure 1:**
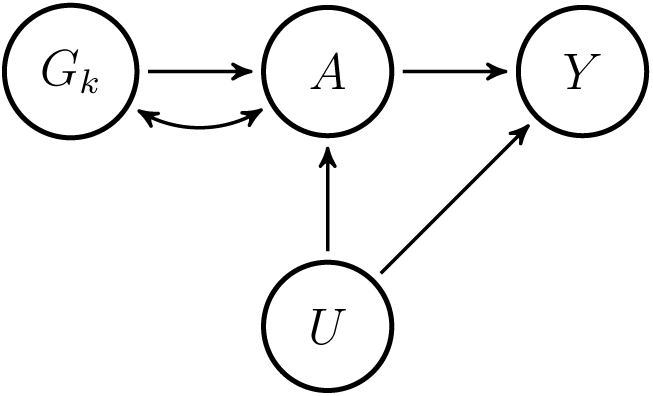
Causal diagram representing the IV model within strata of *𝒢 \G*_*k*_. The bi-directed arrow between *G*_*k*_ and *A* indicates potential unmeasured common causes of *G*_*k*_ and *A*.

### Assumption 4.

There is no additive *A −*(*U*, 𝒢) interaction in model for *E* (*Y* |*A*, 𝒢, *U*)

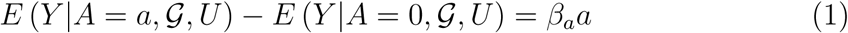

Note that the condition does not require the vector 𝒢 to be a valid IV. Equation (1) implies that the additive causal effect of *A* on *Y* is not modified by either *U* or 𝒢. These restrictions imply the following semi-additive models:

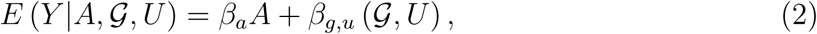

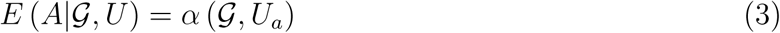

where *β*_*g,u*_ (*·*) and *α*(*·*) are functions solely restricted by natural features of the model, e.g. such that the outcome and exposure means are bounded between zero and one in the binary case. If *G*_*k*_* is a valid IV, then *β*_*g,u*_ (𝒢, *U*) = *β*_*g,u*_ (𝒢 _*−k*_*, *U*) where 𝒢 _*−k*_* = 𝒢 *\G*_*k*_*, by the exclusion restriction implying Assumption 4 reduces to the assumption of no (*U*, 𝒢 _*−k*_*) *−A* interaction in the model for *E* (*Y* |*A*, 𝒢, *U*) ; Furthermore, we have that Pr*{E* (*A*|𝒢, *U*) *−E* (*A*|𝒢 _*−k*_, *G*_*k*_* = 0, *U*) = 0} ≠ 0 by Assumption 1. Note that models *E*(*Y* |*A*, 𝒢, *U*) and *E*(*A*|𝒢, *U*) considered by Bowden et al. (2015) satisfy (2) and(3) with *β*_*g*_ (*·*) and *α* (*·*) linear functions, while Kang et al. (2016) specified models implied by these two restrictions; Tchetgen Tchetgen et al. (2021) considered inference with invalid IVs under model (2) and *E* (*A*|𝒢, *U*) = *α*_*G*_ (𝒢) + *α*_*U*_ (*U*), and Pr {Var (*A*|𝒢) *−*Var (*A*|𝒢 =0) = 0*} ≠* 0. Clearly, the framework we are considering subsumes models considered in these prior works. Despite its widespread use, the semi-additive model can be mis-specified in presence of an additive interaction between treatment and either an invalid IV SNP, or an unmeasured confounder in the outcome model. It may also fail to hold in case of nonlinearity in dose-response curve for a continuous exposure. In both cases the model can be viewed as a first order approximation to the extent that departure from the additive structure is relatively small. Still, it is also important to note that even with an IV known ex ante to be valid, a homogeneity condition akin to (2) for the outcome model or a similar condition for the treatment model appears to be necessary for point identification of causal effects, see Robins (1994), Angrist et al. (1996) and Wang and Tchetgen Tchetgen (2018). Crucially, the semi-additive model is guaranteed to hold under the null hypothesis of no additive causal effect, in which case a distribution-free test of the null becomes feasible. In Section 7, partial identification of average causal effects with many invalid IVs is considered under a fully nonparametric model that does not impose restriction (2).

## 3 Identification for Many Independent Invalid IVs

In this section, we state our results under the following joint independence assumption of genetic variants included in 𝒢, which we later relax.

### Assumption 5.

Let 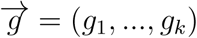 ∈supp(𝒢) then

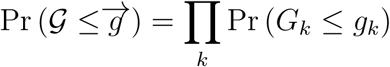

This assumption may be reasonable in case genetic variants encode presence of a minor allele at single polymorphism locations under linkage equilibrium of 𝒢. In practice, if genetic variants are in linkage disequilibrium (LD), then one can perform LD clumping to choose an independent set of genetic variants as candidate instruments using the well-established genetics analysis software PLINK (Purcell et al., 2007).

Next, suppose that unbeknownst to the analyst, Assumptions 1-3 hold for *G*_*k*_*, so that any of the genetic variants in 𝒢 *\G*_*k*_* may be invalid. In order to ease the exposition, we give our first identification result in the special case of binary *G*_*k*_, *k* = 1, …, *K*. For example, *G*_*k*_ may encode whether the risk/protective allele is present at the *k*-th genetic location.

### Proposition 1.

*Suppose that {G*_*k*_ : *k} are binary random variables, and G*_*k*_* *satisfies Assumptions 1-3, furthemore suppose that Assumptions 4 and 5 hold, then*

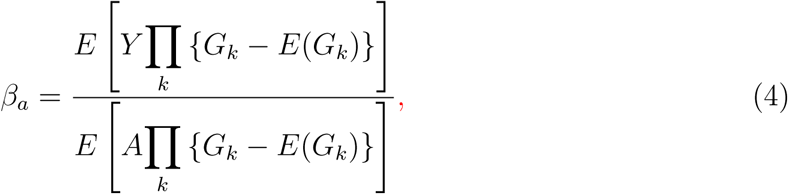

*provided that*

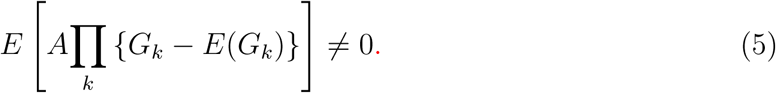

Formal proofs of all results are given in the Supplementary Materials. Proposition 1 provides an explicit identifying expression for the average causal effect *β*_*a*_ of *A* on *Y* in the presence of additive confounding, which leverages the fact that at least one candidate genetic IV *G*_*k*_* is valid, without knowing which are invalid genetic IVs. Notably, the formula (4) given in the Proposition 1 is a natural generalization to accommodate many invalid genetic IVs, of the standard IV estimand which an Oracle with knowledge of *k*^*^ would in principle be able to evaluate in the population:

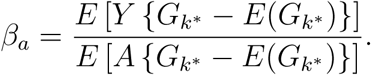

It is instructive to note that the left-hand side of (5) is equivalent to the following being a nontrivial function of all variables in 𝒢 _*−k*_*,

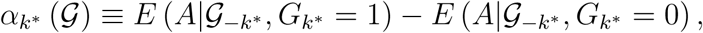

i.e. the *K*^*th*^ order additive interaction of 𝒢 in *E* (*A*|𝒢) is non-null. Intuitively, the Proposition can be motivated upon noting that if *G*_*k*_* is a valid IV, then the main effects and additive interactions of all order involving *G*_*k*_* are excluded from *β*_*g*_ (𝒢) and therefore, the main effect of *G*_*k*_* and all its interactions could in principle be used as potential IVs provided they are not also excluded from the first stage regression function *E* {*α* (𝒢, *U*_*a*_) |𝒢 }. Now, without knowledge of *k*^*^, it is generally not possible to identify ex ante which of these main effects or interactions can be excluded from *β*_*g*_ (𝒢) without error, as we might inadvertendly exclude an invalid IV or interactions of invalid IVs; with the exception of the *K*^*th*^ order additive interaction which is a priori known with certainty to be excluded from *β*_*g*_ (𝒢) when one or more candidate IVs satisfies the exclusion restriction, in which case *α*_*k*_* (𝒢) must depend on the *K*^*th*^ order interaction in order for the latter to be a valid IV, i.e. for IV relevance to also hold. In MR, such higher-order interaction among genetic markers is an important biological phenomenon that has been extensively studied in the genetics literature (Ritchie et al., 2001; Domingo et al., 2018). In order to illustrate the result, suppose that *K* = 2, then, without loss of generality assuming that unbeknownst to the analyst, *G*_1_ is in fact a valid IV, while *G*_2_ is not. Consider the exposure conditional mean *E*(*A*|𝒢) = *α*_0,0_ + *α*_1,0_*G*_1_ + *α*_0,1_*G*_2_ + *α*_1,1_*G*_1_*G*_2_ = (*α*_0,0_ + *α*_0,1_*G*_2_) + *G*_1_(*α*_1,0_ + *α*_1,1_*G*_2_). Then, according to the theorem,

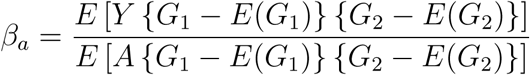

provided that, for binary *G*_1,_*G*_2_ :

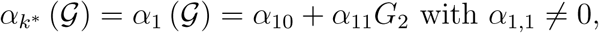

a more stringent IV requirement than standard IV relevance which requires that either *α*_1,0_ ≠ 0 or *α*_1,1_ ≠ 0.

Interestingly, the following result firmly establishes that above strengthening of IV relevance is in fact a necessary condition for the existence of a regular and asymptotically linear (RAL) estimator of *β*_*a*_ under (2) and (3) such that *β*_*g,u*_ (𝒢, *U*) = *β*_*g,u*_ (𝒢 _*−k*_*, *U*) without a priori knowledge of *k*^*^. In this vein, consider the following semiparametric model ℳ_*k*_* characterized by (i) the following observed data restriction implied by structural model (2):

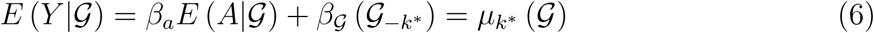

and (ii) Assumption 5. The following result establishes a necessary condition for existence of a RAL estimator under the semiparametric union model 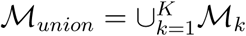 in which at least one SNP is a valid IV without ex ante knowledge which SNPs are invalid IVs.

### Theorem 1.

*Any RAL estimator* 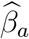 *of β*_*a*_ *in ℳ*_*union*_ *must satisfy the following:*

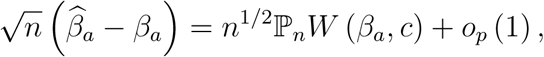

*Where*

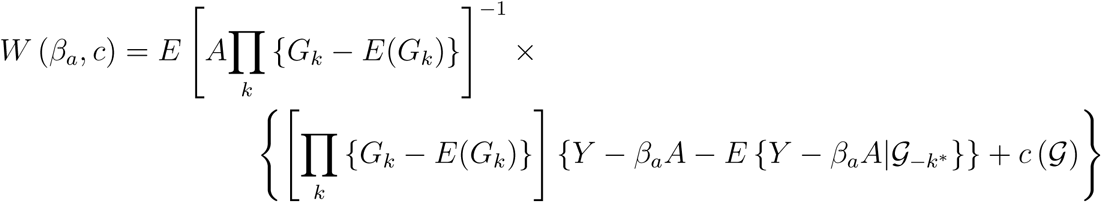

*for some function c* (𝒢) *that satisfies the restriction*

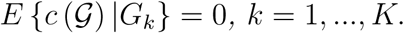

*Furthermore, the efficient influence function of β*_*a*_ *in ℳ*_*union*_ *is given by W* (*β*_*a*_, 0) *and the semiparametric efficiency bound of β*_*a*_ *in ℳ*_*union*_ *is given by*

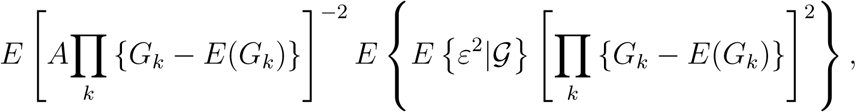

*Where*

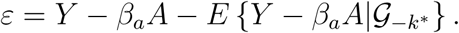

An immediate corollary of the above result is that

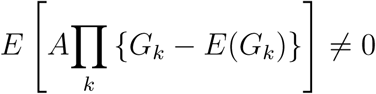

is in fact a necessary condition for the existence of a RAL estimator for *β*_*a*_ in ℳ_*union*_ with a finite asymptotic variance. Because at least 1 out of *K* putative IVs are known *a priori* to be valid, but we do not know the identity of the valid IV 𝒢_*k*_*, the influence function in Theorem 1 may be evaluated as

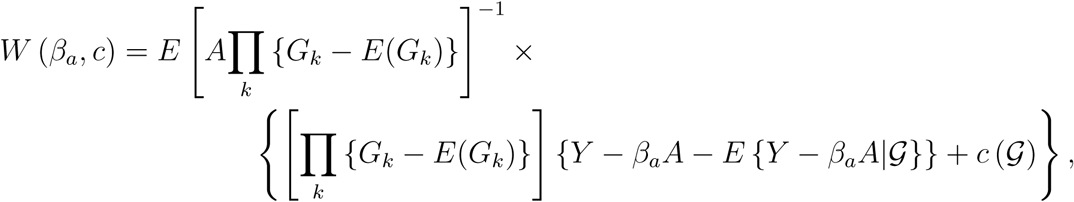

which is an equivalent representation in ℳ_*union*_ due to (6). In the next Section, we establish a more general result by considering a less restrictive invalid IV model in which at least *k*^†^ *>* 1 SNPs are known ex ante to be valid IVs, without knowledge of which SNPs are invalid IVs. This general framework is particularly instructive as it reveals a fundamental trade-off between the strength of required IV relevance condition, and the potential number of invalid IVs one is willing to allow in a given MR analysis

### 3.1 *K* Choose *k*^†^ Multiple Robustness

In this section, we derive estimators for *β*_*a*_ under model (2) provided at least *k*^†^ *>* 1 SNPs are known ex ante to be valid IVs, without necessarily knowing which SNPs are invalid IVs. An important limitation of inference under ℳ_*union*_ is its over reliance on the strong IV relevance requirement, that a *K−*way additive interaction involving all candidate SNP IVs be sufficiently related to the treatment to ensure root-n consistency and asymptotic normality of the estimated treatment effect. Such a requirement may be overly conservative in practice, and prone to weak IV bias if the *K−*way additive interaction in the first stage regression of the endogenous treatment on candidate SNP IVs is weak. Also, it could be that the analyst has a significant degree of ex ante confidence that at least *k*^†^ *>* 1 SNPs are valid IVs; see for instance Kang et al. (2016) and Windmeijer et al. (2019) for methods that assume ex ante that at least 50% of IVs are valid. Alternatively, even when such a priori knowledge is lacking, one may wish to alleviate weak IV concerns, by conducting a sensitivity analysis in which a consistent estimator is obtained assuming at least *k*^†^ valid IVs for *k*^†^ = 1, …, *K*.

The following result generalizes Proposition 1 to this more general setting. Consider for a fixed value of *k*^†^, the semiparametric model 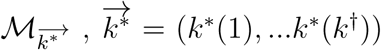 with *k*^*^(1) *<* … < *k*^*^(*k*^†^) ∈ {1, …, *K*} in which Assumption 5 holds, and structural model (2) holds with 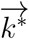 valid IVs, which further implies that:

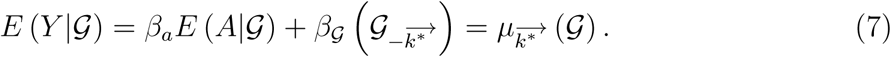

Note that, for notational convenience, we have suppressed dependence on *k*^†^, and *G*_*k*_ may not necessarily be binary. The following result provides an identification result of *β*_*a*_ under the semiparametric union model 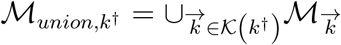 where the union is taken over all elements of

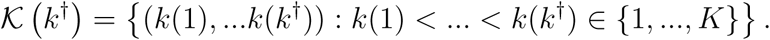

ℳ_*union,k*_^†^ is formally the set of all observed data distributions in which Assumption 5 holds, and model (7) holds for at least *k*^†^ SNPs without ex ante knowledge which SNPs are Invalid IVs among the remaining *K −k*^†^ SNPs.

#### Proposition 2.

*Under model ℳ*_*union,k*_^†^, *we have that:*

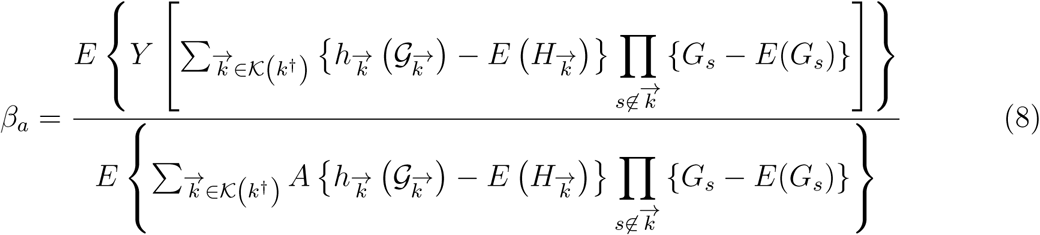

*Provided that*

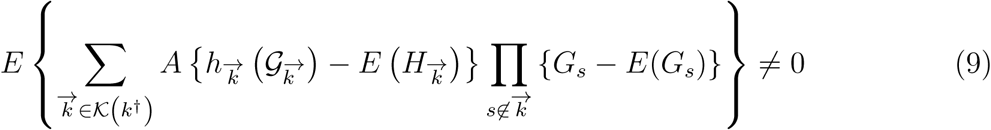

The result provides an explicit expression of the identifying formula for *β*_*a*_ under *M*_*union,k†*_, the union semiparametric model under which at least *k*^†^ out of *K* IVs are known *a priori* to be valid, without knowing which IVs are invalid.

#### Remark 1.

*The multiple robustness result of Proposition 2 is distinct from prior literature which primarily concerns nuisance model specification under a single causal model, whereas our notion of multiply robust inference concerns identification in the union of causal models M*_*union,k*_^†^. *In fact, each model* 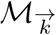 *of the union model of the Proposition may be represented by a distinct causal diagram such as Figure 1. To the best of our knowledge, Proposition 2 constitutes the first instance of causal identification multiple robustness property. In contrast, existing literature on multiple robustness has primarily focused on nuisance model specification under a common set of causal identification, e*.*g. two out of three nuisance model specification robustness in mediation analysis (Tchetgen Tchetgen and Shpitser, 2012) as well as doubly robust estimation of the average treatment effect under unconfoundedness conditions and related missing data problems (Scharfstein et al*., *1999)*.

In order to illustrate the result, suppose that *K* = 5 and *k*^†^ = 2; then, without loss of generality assuming that unbeknownst to the analyst, *G*_1_ and *G*_2_ are in fact valid IVs, while *G*_3_, *G*_4_, *G*_5_ are not, according to the theorem,

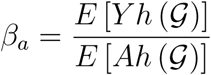

where

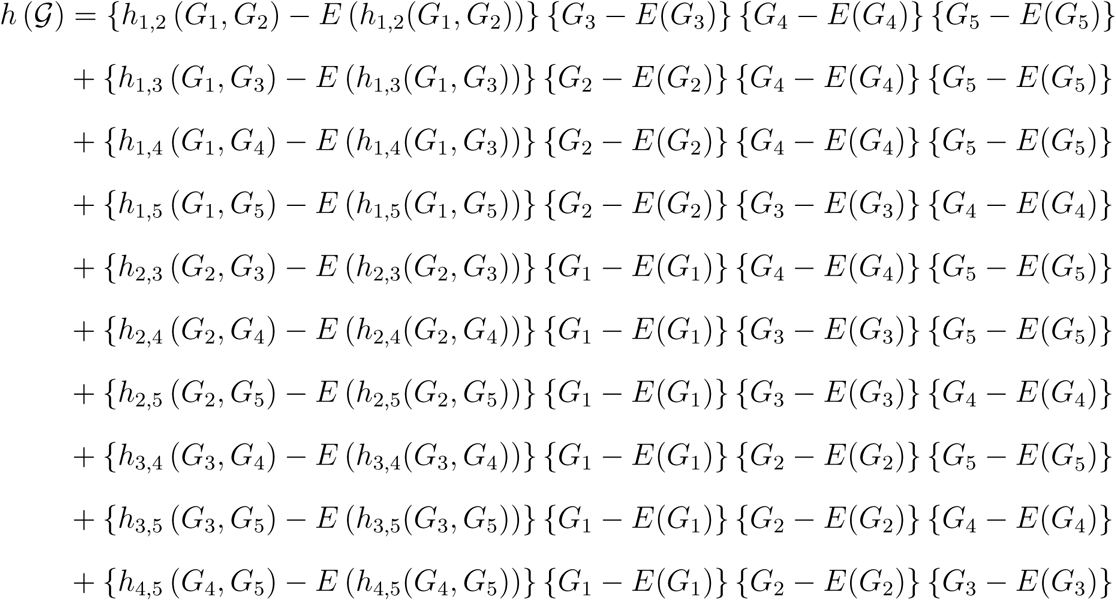

provided that,

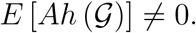

An intuitive interpretation of this identification result is obtained by noting that by virtue of (*G*_1_, *G*_2_) being valid IVs,

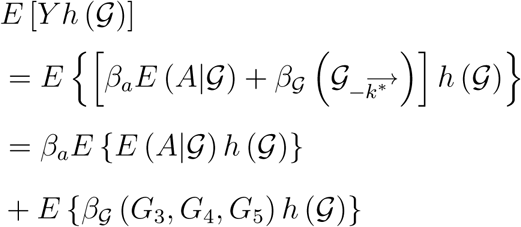

Next, we note that *E* {*h* (𝒢) |*G*_3_, *G*_4_, *G*_5_} = 0 which implies

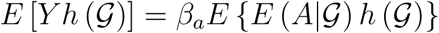

Likewise it is straightforward to verify that

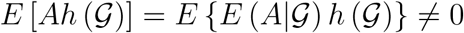

only if *E* (*A*|𝒢) depends on a four-way interaction of form *G*_1_*G*_3_*G*_4_*G*_5_ or *G*_2_*G*_3_*G*_4_*G*_5_; or the five-way interaction *G*_1_*G*_2_*G*_3_*G*_4_*G*_5_. Clearly, this is a weaker requirement than for inference under *K* = 5 and *k*^†^ = 1, which would require that *E* (*A*|𝒢) definitely include the nonzero five-way additive interaction *G*_1_*G*_2_*G*_3_*G*_4_*G*_5_. By the same token, inferences under *K* = 5 and *k*^†^ = 3 can be shown to be possible if say *G*_1_, *G*_2_ and *G*_3_ are in fact valid IVs, while *G*_4_, *G*_5_ are invalid IVs, provided that *E* (*A*|𝒢) depends on a three-way interaction *G*_1_*G*_4_*G*_5_, *G*_2_*G*_4_*G*_5_, or *G*_3_*G*_4_*G*_5_; or alternatively on a four-way interaction *G*_1_*G*_3_*G*_4_*G*_5_, *G*_1_*G*_2_*G*_4_*G*_5_ or *G*_2_*G*_3_*G*_4_*G*_5_; or alternatively on the five-way interaction *G*_1_*G*_2_*G*_3_*G*_4_*G*_5_. This example illustrates that the more candidate SNPs are valid IVs, the less stringent is the IV relevance assumption required for identification. In fact when all five SNPs are valid IVs, then the above result provides an identifying expression for *β*_*a*_ provided at least one of the SNPs is associated with the treatment. We emphasize that there exists strong evidence that genetic higher-order interactions contribute to complex traits (Ritchie et al., 2001; Taylor and Ehrenreich, 2015; Domingo et al., 2018). For example, Ritchie et al. (2001) identified a statistically significant high-order interaction among four SNPs from three different estrogen-metabolism genes in breast cancer studies, even in the absence of any statistically significant main effects. At any rate, appropriateness of the interaction-IVs relevance assumption in question for a given value of *k*^†^ is empirically testable as later illustrated in the UK BioBank Analysis.

We also have the following result establishing a necessary condition for existence of a RAL estimator under ℳ_*union,k*_†.

#### Theorem 2.

*Any RAL estimator* 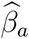 *of β*_*a*_ *in ℳ*_*union,k*_^†^ *must satisfy the following:*

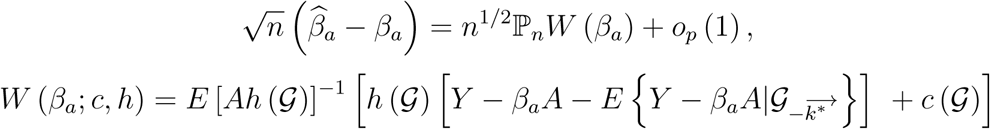

*for a function c* (𝒢) *that satisfies the restriction*

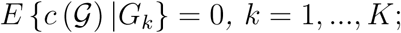

*and functions* 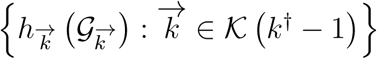 *such that*

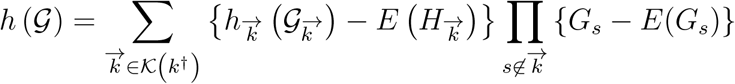

*provided that E* (*A*|𝒢) *depends on at least one nonzero higher order interaction of order greater or equal to K k*^†^ + 1 *involving all invalid IVs* 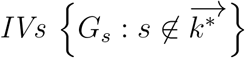 *and at least one valid IV* 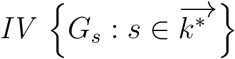. *Furthermore, an estimator which achieves the semiparametric efficiency bound for ℳ*_*union,k*_^†^ admits the above expansion with influence function given by *W* (*β*_*a*_; *c* = 0, *h*_*opt*_), *with h*_*opt*_ *given in the Supplementary Materials*.

### 3.2 MR^2^ Estimation

Theorem 2 provides a novel characterization of all functions that have zero-mean conditional on 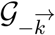 for all 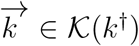 This general result motivates the following MR^2^ estimator via two stage least squares. Specifically, to ground ideas, suppose for each vector 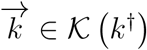, the analyst defines 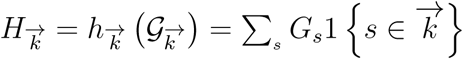, which in case *G*_*s*_ indicates presence of a minor allele at SNP locus *s*, counts the number of SNPs w a minor allele present among all SNPs in 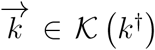. We then note that one can construct valid IVs from *{G*_*s*_ : *s}* under ℳ _*union,k*_^†^ as followed:

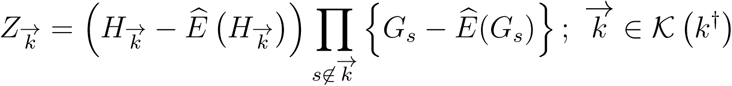

Then, upon obtaining these valid IVs, a standard two stage least squares approach entails:

1. Regressing *A* on valid 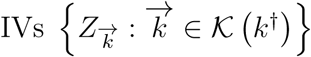 by OLS, with corresponding fitted values

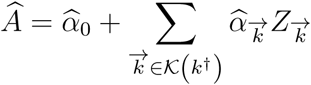
2. Regressing *Y* on *Â* by OLS to obtain 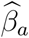.

Then 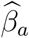 is a consistent and asymptotically normal MR^2^ estimator of *β*_*a*_ under ℳ _*union,k*_^†^, with influence function given in the Supplementary Materials, which can be used for inference, such as to construct 95% confidence intervals for *β*_*a*_. An appealing feature of the MR^2^ estimator is that it is able to attain the semiparametric efficiency bound asymptotically under homoskedasticity *E*(*ε*^2^| 𝒢) = *E*(*ε*^2^) by replacing the original candidate IVs with a corresponding set of generated valid IVs. For improved efficiency, one can further adjust for certain eligible functions of SNPs under ℳ _*union,k*_^†^. Specifically, we note that under ℳ _*union,k*_^†^, higher order interactions of order greater or equal to *K −k*^†^ + 1 comprise valid IVs, however interactions of order strictly lower than *K −k*^†^ + 1 may not be valid IVs as they may be associated with the outcome, in which case they might be of use to reduce variability in the outcome, and thus improve estimation efficiency, and when also associated with the treatment, they might also partially account for confounding.

In this vein, we let *X* denote a vector of eligible covariates, which may include main effects, together with second, third, …. *K −k*^†^ *th* order interactions of *{G*_*s*_ : *s}*. Then, to the extent that they are predictive of the outcome, further adjusting for *X* in stage 2 outcome regression by regressing *Y* on (*Â, X*) can yield a more efficient estimator 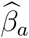 for the regression coefficient of *Â* Further adjusting for other observed covariates, which we may denote as *M* to account for possible confounding of the effect of SNPs on the outcome typically due to population stratification (i.e. confounding by ancestry), can likewise be implemented. This can be done by simply replacing 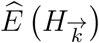 with 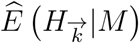 and *Ê*(*G*_*s*_) with *Ê*(*G*_*s*_|*M*) obtained by fitting standard regression models for 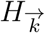 and *G*_*s*_ on *M* respectively, in construction of 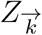, and by including both *X* and *M* in the stage 2 outcome regression model.

## 4 Simulation Study

We perform Monte Carlo simulations to investigate the numerical performance of the proposed estimators. The instruments 𝒢 = (*G*_1_, …, *G*_*K*_)^*T*^ are generated from independent Bernoulli distributions with probability *p* = 0.8. Similar to the study designs in Guo et al. (2018) and Windmeijer et al. (2019), the outcome and exposure are generated without covariates from

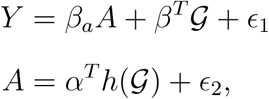

where *α* encodes all main and interaction effects of *𝒢*, and

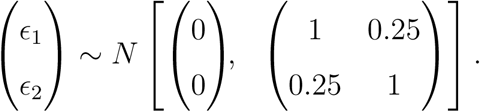

Under this data generating mechanism, we set the number of IVs to be *K* = 5, the causal effect parameter *β*_*a*_ = 1 and vary

1. the sample size *n* = 10000 or 50000;
2. the number of valid IVs *K −*||*β*||_0_; and
3. the IV strength *α* = (1, …, 1)^*T*^ *C* for *C* = 0.6 or 1.

For (2), we first set *β* = (0, 0, 0, 0.2, 0.2)^*T*^ encoding three valid IVs so that the 50% rule of Kang et al. (2016) holds. Then we consider the case where only two IVs are valid, and set *β* = (0, 0, 0.1, 0.2, 0.3)^*T*^ or *β* = (0, 0, 0.2, 0.2, 0.2)^*T*^. The plurality rule of Guo et al. (2018) holds in the former design but is violated in the latter.

We use the R package `AER` (Kleiber and Zeileis, 2008) to implement the semiparametric efficient MR^2^ estimator under homoskedasticity when there are at least 2 valid IVs, i.e. in the union model ℳ _*union*,2_. A sandwich estimate of the asymptotic variance is available directly as an output from the function ivreg. We compare the MR^2^ method with

1. the oracle two stage least squares (2SLS) estimator using only the valid IVs;
2. the naive 2SLS estimator using all of 𝒢;
3. the post-adaptive Lasso (Post-Lasso) estimator of Windmeijer et al. (2019) which uses adaptive Lasso tuned with an initial median estimator to select valid IVs; and
4. the two-stage hard thresholding (TSHT) estimator of Guo et al. (2018).

The oracle 2SLS estimator is included as a benchmark. It requires *a priori* knowledge of which of the putative IVs are valid, and thus is infeasible in practice. Table 1 summarizes the estimation results for the setting with *C* = 0.6 based on 1000 replications. When the 50% rule holds, both Post-Lasso and TSHT perform nearly as well as the oracle 2SLS in terms of absolute bias, coverage and variance. When the 50% rule is violated but the plurality rule holds, TSHT performs as well as the oracle 2SLS once *n* = 50000. As Post-Lasso relies on the 50% rule condition for consistency, it performs poorly in this setting. When both the 50% and the plurality rules are violated, Post-Lasso and TSHT exhibit noticeable bias and poor coverage. In agreement with theory, naive 2SLS suffers from poor coverage throughout, regardless of sample size.

**Table 1:** Comparison of methods with continuous exposure generated under the identity link (*C* = 0.6). The two rows of results for each estimator correspond to sample sizes of *n* = 10000 and *n* = 50000 respectively.

|  | MR<sup>2</sup> | Oracle 2SLS | Naive 2SLS | Post-Lasso | TSHT |
| --- | --- | --- | --- | --- | --- |
| 50% rule holds |  |  |  |  |  |
| \|Bias\| | .007 | .000 | .014 | .000 | .000 |
|  | .002 | .000 | .014 | .000 | .000 |
| $\sqrt{\text{Var}}$ | .019 | .002 | .002 | .003 | .003 |
|  | .009 | .001 | .001 | .001 | .002 |
| $\sqrt{\text{EVar}}$ | .019 | .002 | .002 | .003 | .003 |
|  | .009 | .001 | .001 | .001 | .001 |
| Cov95 | .929 | .951 | .000 | .940 | .950 |
|  | .943 | .945 | .000 | .947 | .947 |
| 50% rule violated but plurality rule holds |  |  |  |  |  |
| \|Bias\| | .032 | .000 | .021 | .014 | .009 |
|  | .017 | .000 | .021 | .012 | .000 |
| $\sqrt{\text{Var}}$ | .058 | .003 | .002 | .009 | .009 |
|  | .043 | .001 | .001 | .007 | .003 |
| $\sqrt{\text{EVar}}$ | .068 | .003 | .002 | .003 | .003 |
|  | .047 | .001 | .001 | .001 | .001 |
| Cov95 | .935 | .938 | .000 | .118 | .424 |
|  | .933 | .951 | .000 | .000 | .944 |
| Both 50% and plurality rules violated |  |  |  |  |  |
| \|Bias\| | .032 | .000 | .021 | .035 | .035 |
|  | .017 | .000 | .021 | .036 | .035 |
| $\sqrt{\text{Var}}$ | .057 | .003 | .002 | .003 | .003 |
|  | .043 | .001 | .001 | .001 | .002 |
| $\sqrt{\text{EVar}}$ | .068 | .003 | .002 | .002 | .002 |
|  | .047 | .001 | .001 | .001 | .001 |
| Cov95 | .933 | .941 | .000 | .000 | .000 |
|  | .933 | .950 | .000 | .000 | .000 |

Note: |Bias| and $\sqrt{\text{Var}}$ are the Monte Carlo absolute bias and standard deviation of the point estimates, $\sqrt{\text{EVar}}$ is the square root of the mean of the variance estimates and Cov95 is the coverage proportion of the 95% confidence intervals, based on 1000 repeated simulations. Zeros denote values smaller than .0005.

The proposed MR^2^ estimator performs well across all simulation settings, including those in which both the 50% and the plurality rules are violated, but with substantially higher variance than the other methods. For inference, the standard sandwich estimator of the asymptotic variance which ignores data-dependency of the estimated means {*Ê* (*G*_*j*_)}_*j*=1,…,*K*_ is conservative (Robins et al., 1994), albeit readily available as an output from existing off-the-shelf software. As shown in the Supplementary Materials, the degree of conservativeness increases with the amount of outcome variation that can be explained by the IVs. Thus when there are more invalid IVs having direct effects on the outcome, e.g. in settings with 50% rule violated in Table 1, the conservativeness is more pronounced. A convenient alternative for obtaining standard errors is to implement the nonparametric bootstrap which appropriately accounts for all sources of uncertainty.

Overall, the simulation study shows that Post-Lasso and TSHT perform better than MR^2^ in terms of efficiency when their respective identifying conditions hold. On the other hand, MR^2^ is able to deliver the desired level of coverage across the range of simulation settings considered. As expected, such robustness comes at the expense of wider confidence intervals, as measured by the square root of the average of squared standard error estimates, and requiring more samples. The absolute bias and variance of the estimators are generally smaller with stronger IV when *C* = 1; because the conclusions are otherwise qualitatively similar, the estimation results for this setting are included in the Supplementary Materials.

### 4.1 Varying Degree of Exposure Interactions

It is also of interest to investigate the performance of estimators under settings where the exposure model is sparse. To mimick the degree of genetic interactions, in addition to the main effects we randomly select *γ* = 30% or 60% of both the lower and higher order interactions (of order *≥* ||*β*||_0_ +1) to have nonzero coefficients in *α*. However, this sampling scheme may render the plurality rule invalid even when *β* = (0, 0, 0.1, 0.2, 0.3)^*T*^, due to potential asymmetric interactions in the exposure model. Therefore, Table 2 summarizes the estimation results with *C* = 0.6 corresponding to the two cases *β* = (0, 0, 0, 0.2, 0.2)^*T*^ and *β* = (0, 0, 0.2, 0.2, 0.2)^*T*^ only, the latter of which represents violation of both the 50% and plurality rules. As expected the proposed MR^2^ has smaller absolute bias and lower variance as the degree of interactions in the exposure model increases, with empirical coverage close to the nominal level throughout. The estimation results when *C* = 1 are reported in the Supplementary Materials, and yield qualitatively similar conclusions.

**Table 2:**
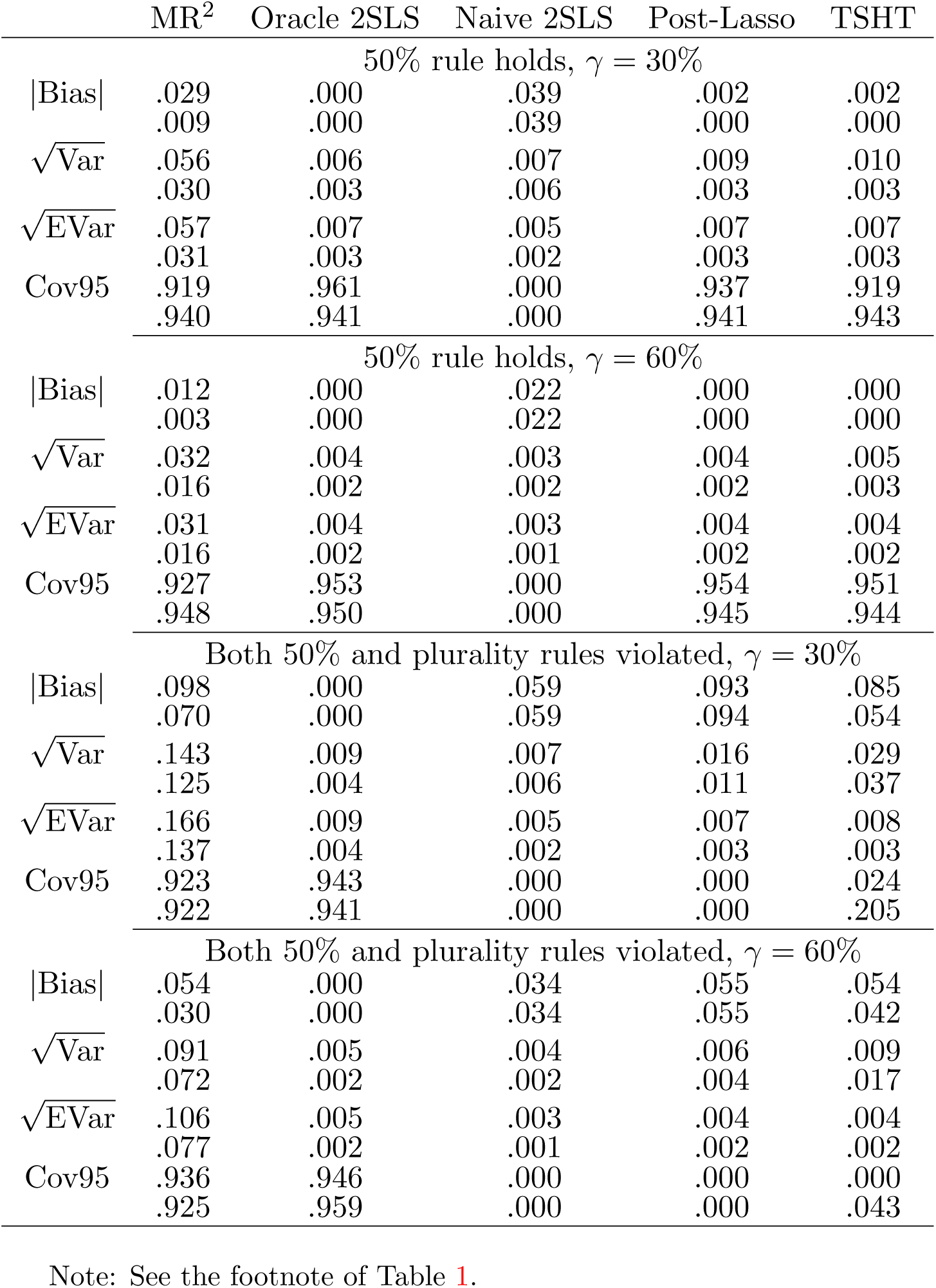
Comparison of methods with continuous exposure generated under the identity link (*C* = 0.6), and varying degree of interactive effects. The two rows of results for each estimator correspond to sample sizes of *n* = 10000 and *n* = 50000 respectively.

### 4.2 Single Index Models

We present further Monte Carlo results for exposures generated under the class of semi-parametric single index models *E*(*A*| 𝒢) = *g*(*α*^*T*^ 𝒢) where *g*(*·*) is a link function. Specifically, the continuous exposure is generated without covariates from

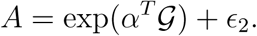

We also consider the important setting of a binary exposure generated from

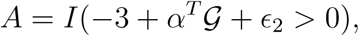

where *I*(*·*) is the indicator function. We fix *C* = 1, while all other simulation settings are unchanged. Tables 3 and 4 summarize the estimation results based on 1000 replications. Remarkably, the proposed MR^2^ method performs well, even though in both settings only main effects are included in the linear predictor function, as exposure interactive effects are induced on the additive scale by the nonlinear link function, rather than specified explicitly *a priori*. All other conclusions are qualitatively similar to those drawn when the exposure is generated under the identity link with interactions.

**Table 3:** Comparison of methods with continuous exposure generated under the log link. The two rows of results for each estimator correspond to sample sizes of *n* = 10000 and *n* = 50000 respectively.

|  | MR<sup>2</sup> | Oracle 2SLS | Naive 2SLS | Post-Lasso | TSHT |
| --- | --- | --- | --- | --- | --- |
| 50% rule holds |  |  |  |  |  |
| \|Bias\| | .000 | .000 | .002 | .000 | .000 |
|  | .000 | .000 | .002 | .000 | .000 |
| $\sqrt{\text{Var}}$ | .001 | .000 | .000 | .000 | .000 |
|  | .001 | .000 | .000 | .000 | .000 |
| $\sqrt{\text{EVar}}$ | .001 | .000 | .000 | .000 | .000 |
|  | .001 | .000 | .000 | .000 | .000 |
| Cov95 | .939 | .951 | .000 | .938 | .950 |
|  | .948 | .945 | .000 | .947 | .946 |
| 50% rule violated but plurality rule holds |  |  |  |  |  |
| \|Bias\| | .002 | .000 | .003 | .002 | .001 |
|  | .000 | .000 | .003 | .001 | .000 |
| $\sqrt{\text{Var}}$ | .006 | .000 | .000 | .001 | .001 |
|  | .003 | .000 | .000 | .001 | .000 |
| $\sqrt{\text{EVar}}$ | .006 | .000 | .000 | .000 | .000 |
|  | .003 | .000 | .000 | .000 | .000 |
| Cov95 | .944 | .939 | .000 | .118 | .423 |
|  | .956 | .951 | .000 | .000 | .944 |
| Both 50% and plurality rules violated |  |  |  |  |  |
| \|Bias\| | .002 | .000 | .003 | .004 | .004 |
|  | .000 | .000 | .003 | .004 | .004 |
| $\sqrt{\text{Var}}$ | .006 | .000 | .000 | .000 | .000 |
|  | .003 | .000 | .000 | .000 | .000 |
| $\sqrt{\text{EVar}}$ | .006 | .000 | .000 | .000 | .000 |
|  | .003 | .000 | .000 | .000 | .000 |
| Cov95 | .940 | .939 | .000 | .000 | .000 |
|  | .956 | .950 | .000 | .000 | .000 |

Note: See the footnote of Table 1.

**Table 4:**
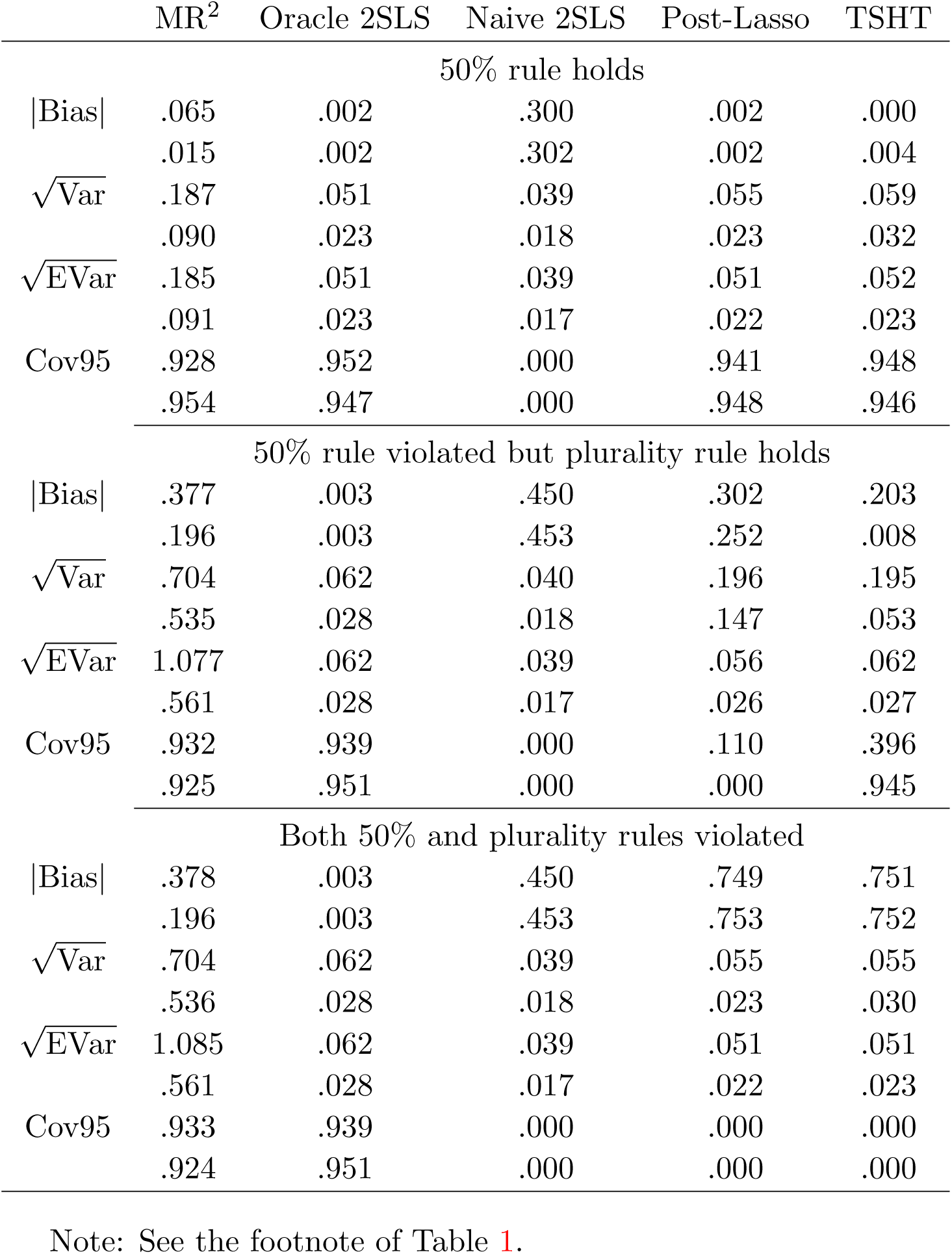
Comparison of methods with binary exposure generated under the probit link. The two rows of results for each estimator correspond to sample sizes of *n* = 10000 and *n* = 50000 respectively.

## 5 An Application to the UK Biobank Data

The UK Biobank (UKB) is a large-scale ongoing prospective cohort study that recruited around 500,000 participants aged 40-69 at recruitment from 2006 to 2010. Participants provided biological samples, completed questionnaires, underwent assessments, and had nurse led interviews. Follow up is chiefly through cohort-wide linkages to National Health Service data, including electronic, coded death certificate, hospital, and primary care data (Sudlow et al., 2015). Genotyping was performed using two arrays, the Affymetrix UK BiLEVE (UK Biobank Lung Exome Variant Evaluation) Axiom array (about 50,000 participants) and Affymetrix UK Biobank Axiom array (about 450,000 participants). The SNPs included for analysis were directly genotyped or imputed using the Haplotype Reference Consortium panel. To reduce confounding bias due to population stratification, we restrict our analysis to people of genetically verified white British descent, as in previous studies (Tyrrell et al., 2016). For quality control, we exclude participants with (1) excess relatedness (more than 10 putative third-degree relatives), or (2) mismatched information on sex between genotyping and self-report, or (3) sex-chromosomes not XX or XY, or (4) poor-quality genotyping based on heterozygosity and missing rates *>* 2%.

We are interested in estimating the causal effects of body mass index (BMI) on systolic blood pressure (SBP) and diastolic blood pressure (DBP) respectively. We exclude participants who are taking anti-hypertensive medication based on self-report. In total, the sample size for the final analysis is 292,757. Among the study samples, the mean (SD) BMI was 27.27 (4.65) *kg/m*^2^, the mean (SD) SBP was 139.6 (19.56) mm Hg, and the mean (SD) diastolic BP was 82.06 (10.66) mm Hg. We use top 10 independent SNPs that are strongly associated with BMI at genome-wide significance level *p* < 5 × 10^*−*8^ (Locke et al., 2015), more detailed information about those 10 SNPs is provided in the Table S1 in the Supplementary Materials. We first perform a conventional regression analysis with BMI as the exposure, SBP/DBP as the outcomes and adjust for sex and age. We obtain a point estimate of 0.680 (SE: 0.007) for SBP and 0.602 (SE:0.004) for DBP for one unit change of BMI. We then apply our proposed MR^2^ method along with the following competing methods: Naive 2SLS, TSHT (Guo et al., 2018), sisVIVE (Kang et al., 2016) and Post-Lasso (Windmeijer et al., 2019).

The data analysis results are summarized in Table 5; note that sisVIVE method does not provide SE estimates. For the BMI-SBP relationship, all effect estimates using MR methods were smaller than estimates from conventional regression analysis. TSHT selected all 10 SNPs as valid IVs, which explains why results are identical for Naive 2SLS and TSHT. Setting *k*^†^ = 10, we obtained a point estimate 0.496 with SE: 0.090. As expected, this was close to naive 2SLS and TSHT results.

**Table 5:** Estimates of the causal effects of body mass index on systolic blood pressure and diastolic blood pressure in the UK Biobank of European descent (*K* = 10, *n* = 292, 757).

| Naive 2SLS | TSHT | sisVIVE | Post-Lasso | MR<sup>2</sup> ($k^\dagger$) | | | |
| --- | --- | --- | --- | --- | --- | --- | --- |
|  |  |  |  | 4* | 6* | 8* | 10* |
| Systolic blood pressure |  |  |  |  |  |  |  |
| .447 | .447 | .604 | .628 | .543 | .523 | .530 | .496 |
| (.109) | (.109) |  | (.133) | (.296) | (.135) | (.090) | (.090) |
| Diastolic blood pressure |  |  |  |  |  |  |  |
| .254 | .296 | .386 | .459 | .649 | .410 | .363 | .349 |
| (.059) | (.059) | — | (.065) | (.147) | (.071) | (.048) | (.048) |

Note: Point estimate with standard error in parenthesis. \*indicates that the weak IV null hypothesis is rejected at 0.05 level using the diagnostic test implemented in the R package **AER** (Kleiberg and Zeileis, 2008).

For the BMI-DBP relationship, TSHT selected nine SNPs as valid, which explains why point estimates for THST and Naive 2SLS differ slightly from each other. Setting *k*^†^ = 10 gave a point estimate of 0.349 with SE: 0.049. Interestingly, the proposed MR^2^ method delivered a point estimate of 0.649 when *k*^†^ = 4, which is larger than those of competing methods. When *k*^†^ = 6 or 8 and thus more than half of the SNPs are assumed to be valid, the point estimate of MR^2^ is similar to those of sisVIVE and Post-Lasso, both of which require the 50% rule for consistency. We also observe that the standard errors of the MR^2^ method decrease substantially when the value of *k*^†^ increases, which is in line with our theory. In particular, when *k*^†^ = 2, the p-value (0.47) for the weak IV diagnostic test in the first stage regression (computed using the R package AER) is not significant, and hence the point estimates are likely to be biased due to weak IVs. For this reason, we do not report results for *k*^†^ = 2. We further perform the Hausman homogeneity test (Hausman, 1978) to compare the point estimates under *k*^†^ = 6, 8, 10 versus that under *k*^†^ = 4 to see whether they differ beyond sampling error using the following test statistic *ht*

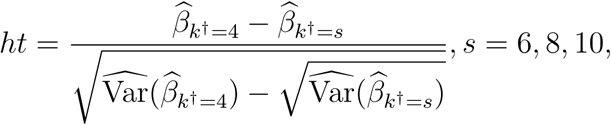

which follows *N* (0, 1) under the null hypothesis *β*_*k*†=4_ = *β*_*k*†=*s*_, *s* = 6, 8, 10, respectively. A large absolute value of the test statistic *ht* suggests that estimators 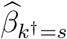 and 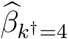 have different limiting values. For the BMI-SBP relationship, we did not find statistically significant differences at 5% level when comparing point estimates for *k*^†^ = 6, 8, 10 with the point estimate for *k*^†^ = 4, indicating that results are relatively insensitive to the assumed number of valid IVs, although inferences for *k*^†^ = 6, 8 may be most reliable. For the BMI-DBP relationship, we found that the point estimate under *k*^†^ = 6 is not statistically different (*p* = 0.063) from that under *k*^†^ = 4, however, point estimates under *k*^†^ = 8, 10 are statistically different (*p <* 0.05) from that for *k*^†^ = 4, which in this case may be most reliable.

## 6 Correlated Invalid IVs

We consider a generalization of the results from Section 3 to allow for correlated SNPs, i.e. without Assumption 5. In context of invalid IVs, this generalization is particularly important for the approach to allow for non-genetic candidate IVs. Consider the semiparametric model which solely asssumes that (7) holds for at least *k*^†^ SNPs without ex ante knowledge of which SNPs are invalid IVs among the remaining *K −k*^†^ SNPs. We have the following result.

### Proposition 3.

*Under model ℳ* _*union,k*_†, *we have that:*

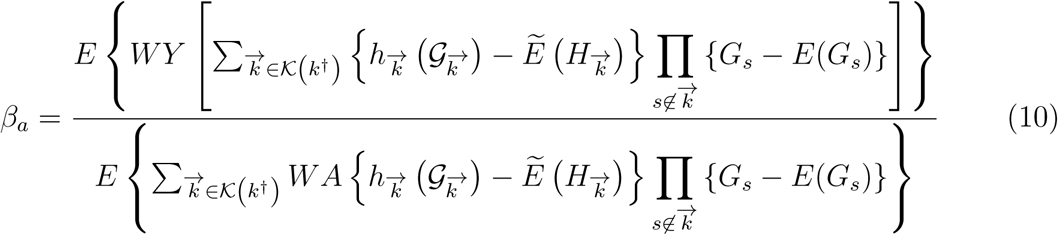

*provided that*

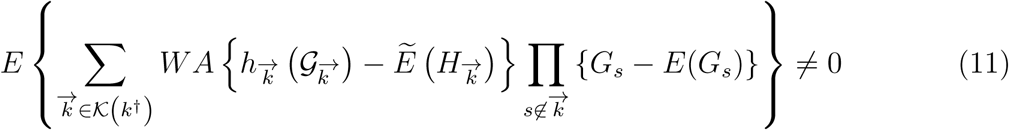

*Where*

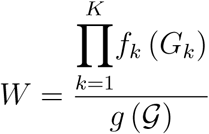

*with g* (𝒢) *the joint probability mass function of* 𝒢 *and f*_*k*_ (*G*_*k*_) *the marginal probability mass function of G*_*k*_, *and the expectation* 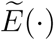 *is taken with respect to the joint probability mass function* 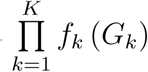 *under independence*.

The weight *W* formally accounts for possible SNP dependence and in fact, we note that under Assumption 5, *W* = 1 for all units, recovering the result from Section 3.

## 7 Partial Identification

The proposed approach also allows for partial identification of average causal effects with many possibly invalid IVs, even in the absence of semi-additive structural restrictions. The key observation that allows one to obtain valid bounds for a causal effect of interest assuming *k*^†^ out of *K* candidate IVs are valid is that, as pointed out in Section 3, such assumption logically implies that all higher order interactions involving at least *K −k*^†^ + 1 candidate IVs must in fact satisfy the exclusion restriction, since such interactions can at most involve *K −k*^†^ invalid IV and at least one valid IV. To illustrate, suppose that *K* = 5 and *k*^†^ = 2; then all four-way interactions *G*_1_*G*_2_*G*_3_*G*_4_, *G*_1_*G*_2_*G*_3_*G*_5_, *G*_1_*G*_2_*G*_4_*G*_5_, *G*_1_*G*_3_*G*_4_*G*_5_, *G*_2_*G*_3_*G*_4_*G*_5_, and the five way interaction *G*_1_*G*_2_*G*_3_*G*_4_*G*_5_ must satisfy the exclusion restriction condition, regardless of which two IVs are valid IVs, because at least one such valid IV is necessarily included in all of these interactions. With these valid candidate interaction-IVs defined, one may then proceed with any existing IV bounds in the literature to obtain partial inference for a variety of average causal effects (Swanson et al., 2018).

## 8 Discussion

In this paper, we introduce a new class of MR^2^ estimators that are guaranteed to be root-*n* consistent for the causal effect of interest in MR studies, provided at least one genetic IV is valid, but without necessarily knowing which one. We further extend the result to a more general class of estimators that remain root-*n* consistent provided that *k*^†^ of the *K* putative IVs are valid. In practice, if one has some prior knowledge about *k*^†^, then such prior knowledge can be incorporated into the analysis. In practice, we propose that one report inferences corresponding to a range of values of *k*^†^ to assess the extent to which inferences vary with more or less stringent IV conditions in a manner of sensitivity analysis. For each value of *k*^†^, one may diagnose weak IVs by inspecting the significance value of the F-statistic in our first-stage regression for the candidate genetic interactions. Both simulation studies and real data analysis demonstrate the finite sample performance of the proposed approach.

Our contribution is multi-fold. First, we by-pass the model selection procedure of Kang et al. (2016); Guo et al. (2018) and Windmeijer et al. (2019); our confidence intervals are based on a regular and asymptotically linear estimator and therefore are uniformly valid over the parameter space of the union model. Second, we establish the semiparametric efficiency bound for estimation of causal effects under a general, union invalid IV model that subsumes models considered by prior works, and provide a formal quantification of the IV relevance requirements and efficiency in exchange for increased robustness. Lastly, we propose MR^2^ two-stage least squares estimators that can be readily implemented using existing off-the-shelf software.

MR^2^ readily extends to binary, count and censored survival outcome settings simply by re-coding the candidate IVs as discussed in Section 7 and using the derived interaction-IVs as valid IVs in appropriate IV analysis for the given type of outcome available in the causal inference literature (Robins, 1994; Abadie, 2003; Vansteelandt and Goetghebeur, 2003; Tan, 2010; Tchetgen Tchetgen et al., 2015; Wang and Tchetgen Tchetgen, 2018; Liu et al., 2020a).

In closing, we acknowledge certain limitations of the MR^2^ method. First, the approach may be vulnerable to weak IV bias which may occur if exposure is weakly dependent on interaction-IVs. However, although not pursued here, in principle, one can leverage existing literature on many weak IVs to address this challenge (Newey and Windmeijer, 2009; Ye et al., 2021). Second, in observational studies the IV assumptions may only hold conditional on a high dimensional set of baseline covariates and the multiple candidate IVs may be dependent, thus requiring methods briefly introduced in Section 6. In such setting, the need to model the joint density of candidate IVs presents several computational and inferential challenges we plan to address in future work.

## Data Availability

The data underlying this article are from the open-access UK Biobank.

https://github.com/zhonghualiu/MRSquared

## Acknowledgment

Eric Tchetgen Tchetgen’s work is funded by NIH grant R01AI104459. Baoluo Sun is supported by the National University of Singapore Start-Up Grant (R-155-000-203-133). This research has been conducted using the UK Biobank resource (https://www.ukbiobank.ac.uk) under application number 44430.

## Supplementary Materials

With a slight abuse of notation, throughout we let 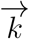 denote either the tuple (*k*(1), …, *k*(*k*^†^)) or the set {*k*(1), …, *k*(*k*^†^)} depending on the context. The complement of set 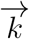 is denoted as 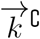, and the set difference 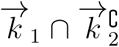 is denoted as 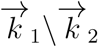 We use 𝒢_*𝒜*_ to denote the set of variables *{G*_*s*_ : *s* ∈ *𝒜 }* for any set of indices *𝒜* ⊆ {1, …, *K}*. In addition, we denote 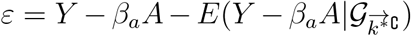 and 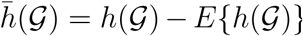 for any function *h*(𝒢). Proposition 1 and Theorem 1 are special cases of Proposition 2 and Theorem 2 respectively with *k*^†^ = 1; their proofs are thus omitted.

## Proof of Proposition 2

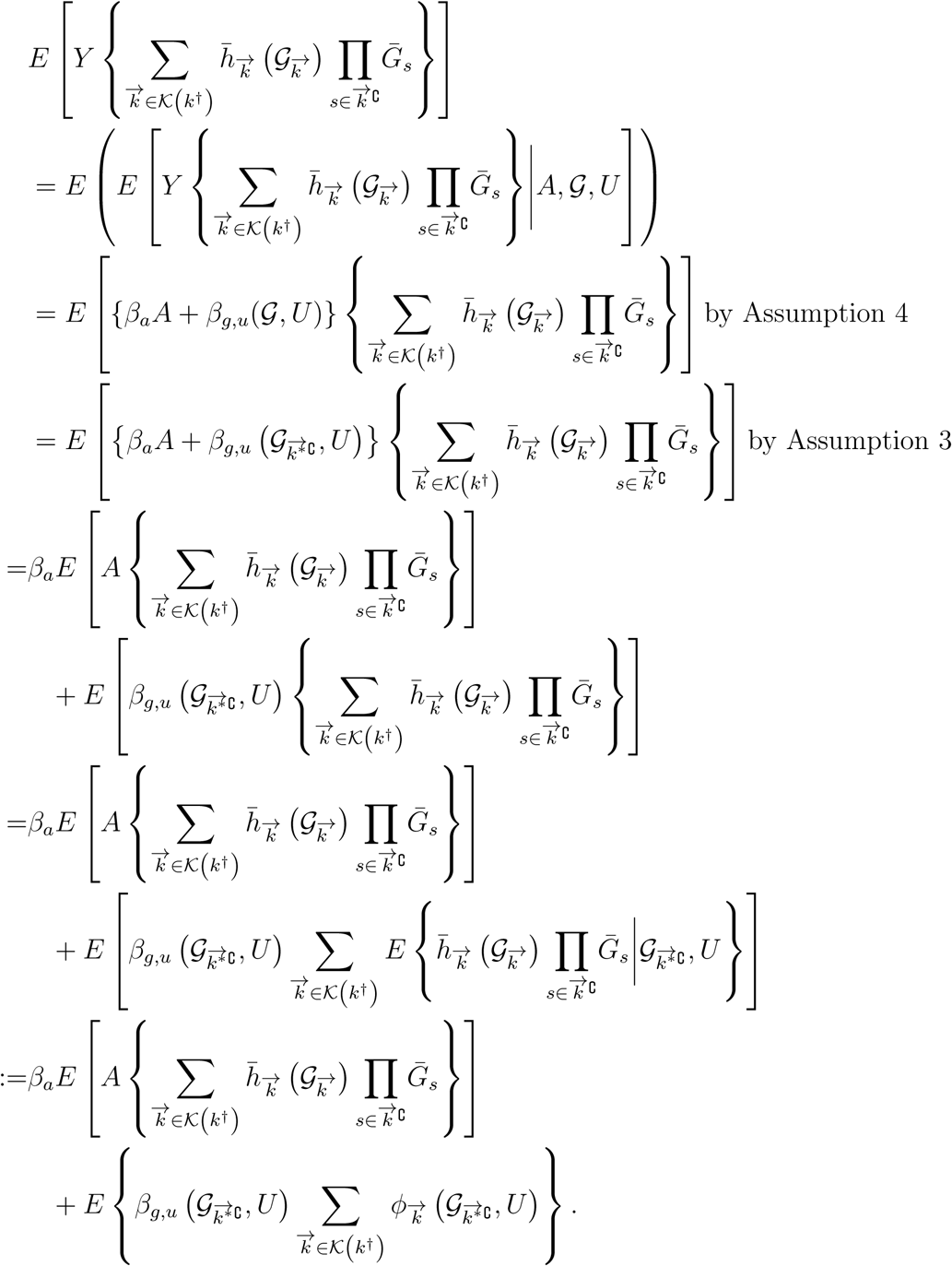

Consider each term 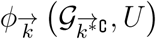, If 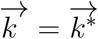, then

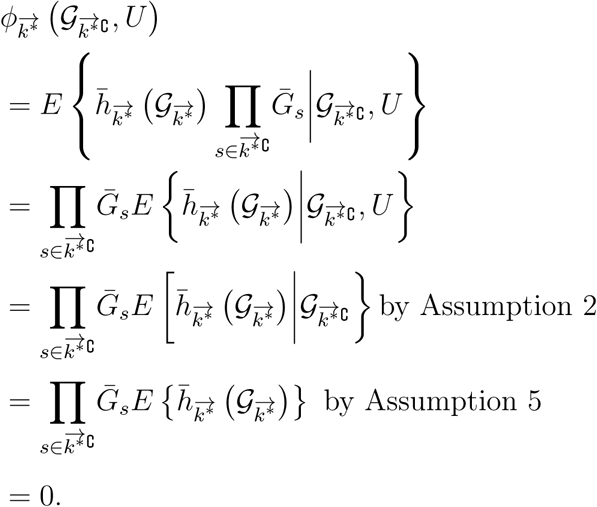

Otherwise 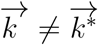, and

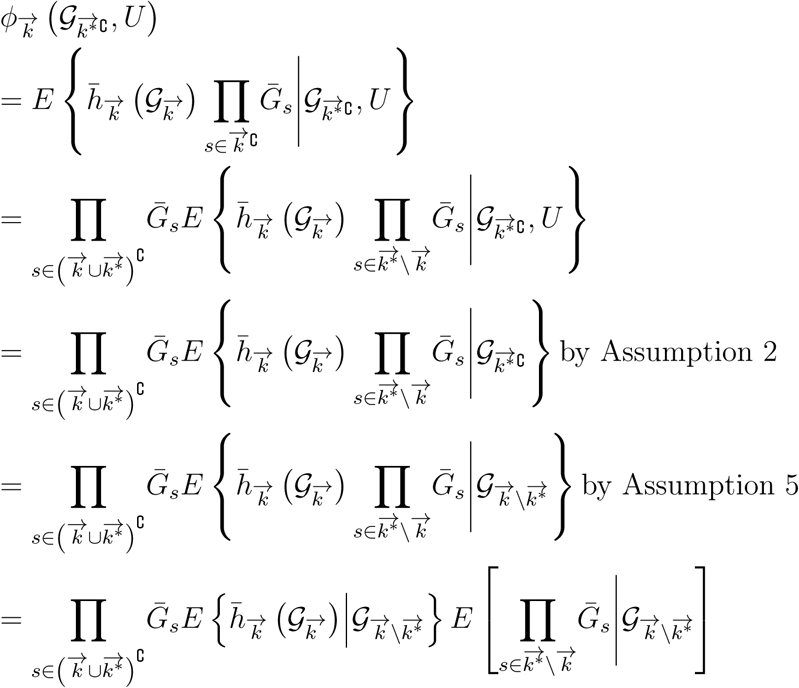

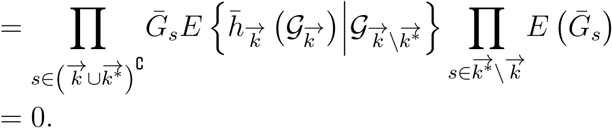

Therefore 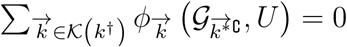 almost surely,and

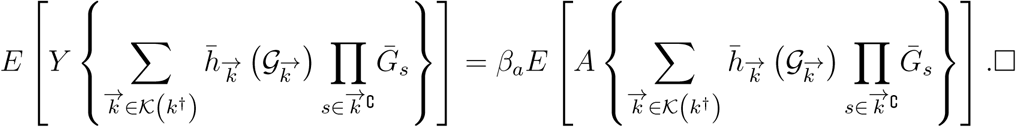

## Proof of Theorem 2

Consider a parametric path *θ* for the joint law of *O* = (*Y, A, G*). The joint density with respect to some dominating measure is given by 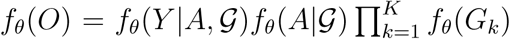 The resulting tangent space is given by 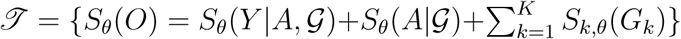., where *E*_*θ*_*{S*_*θ*_(*Y* |*A, 𝒢*)|*A, 𝒢*} = *E*_*θ*_*{S*_*θ*_(*A*| *𝒢*)| *𝒢*} = 0 and *E*_*θ*_*{S*_*k,θ*_(*G*_*k*_)} = 0 for *k* = 1, …, *K*. The parameter of interest *β*_*a*_(*θ*) is defined implicitly as a functional of the observed data law through the conditional mean independence restriction

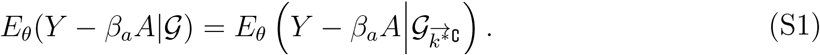

Following Bickel et al. (1993) and Newey (1994), we derive all influence curves IF(*O*) that satisfy

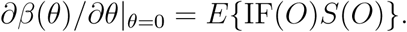

Moment condition (S1) is equivalent to the class of unconditional moment restrictions

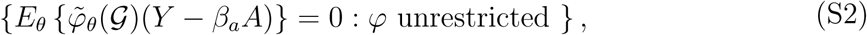

where 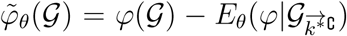 For an arbitrary moment restriction in (S2), differentiating under the integral yields

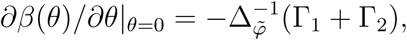

Where 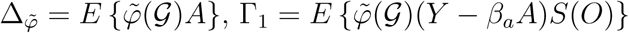 and

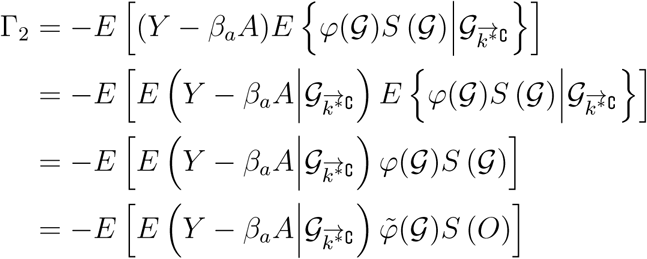

Therefore, 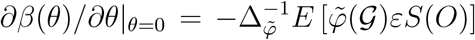 It is straightforward to verify that the orthocomplement to the tangent space is given by

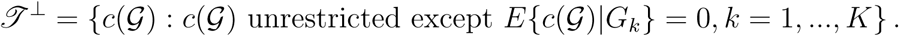

We can add to each influence curve any element from 𝒥 ^*⊥*^, hence the set of influence curves of *β*_*a*_ in semiparametric model 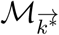 is

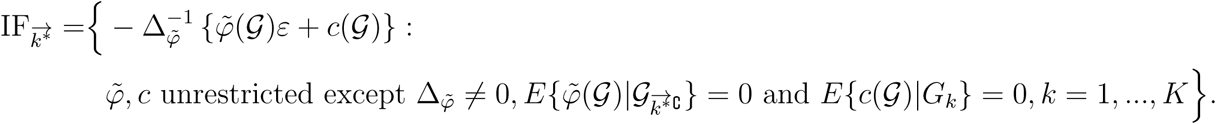

## The Set of Influence Functions in the Semiparametric Union Model *ℳ* _*union,k†*_

By definition, an estimator of *β*_*a*_ is RAL under the union model ℳ _*union,k*_† if and only if it is RAL under each of 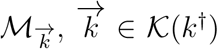, i.e. its influence function is an element in the intersection set

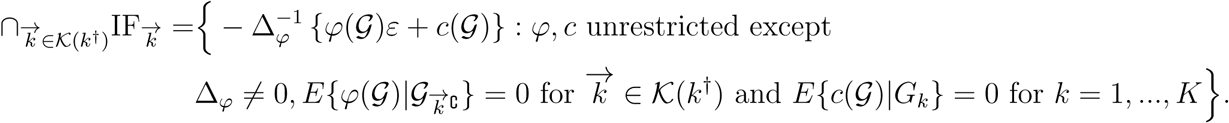

In the following, we consider the enumeration 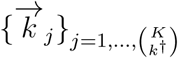 of the elements in *K*(*k*^†^) in Gray code order via the revolving door algorithm (Nijenhuis and Wilf, 2014), which establishes a bijection from the set of integers 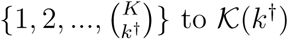 to 𝒦 (*k*^†^) such that 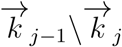 and 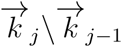 are singletons for any 2 *≤ j ≤ K*. Let 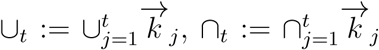 and define

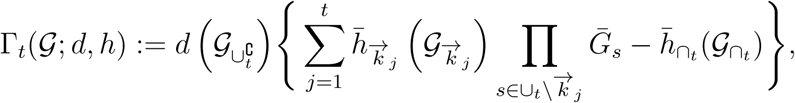

for all 2 *≤ t ≤ K*, where 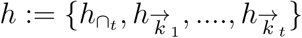 Following the proof of Proposition 3, Γ_*t*_ satisfies

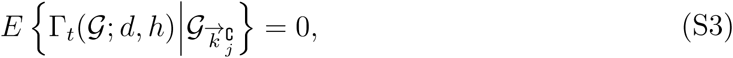

for *j* = 1,, *t*. We also define the set

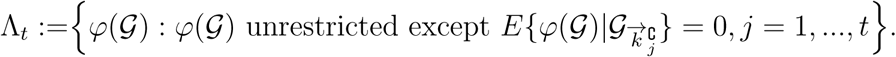

In the following we aim to show that for all 2 *≤ t ≤ K*,

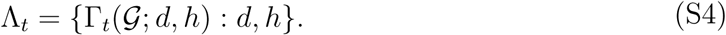

**Proof of** *P* (2): Λ_2_ = {Γ_2_(𝒢; *d, h*) : *d, h*}

The conditional independence 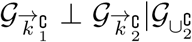 holds under Assumption 5. Then Theorem 1 of Tchetgen Tchetgen et al. (2010) implies that

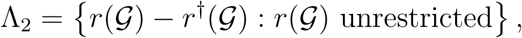

where 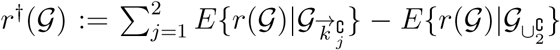 Because 𝒢 consists of all binary variables, any arbitrary function of 𝒢 may be decomposed^1^ as

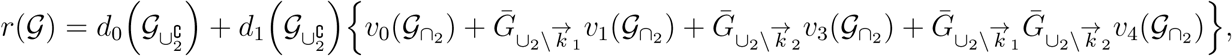

for some functions {*d*_0_, *d*_1_, *v*_0_, *v*_1_, *v*_2_, *v*_3_, *v*_4_}. It may be shown after some algebraic manipulations that

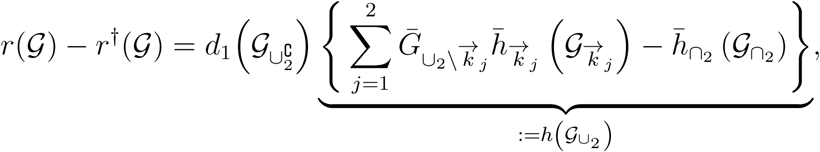

for arbitrary functions *d*_1_ and *h*.

**Proof of** *P* (𝓁) *→ P* (𝓁 + 1) **for** 𝓁 *≥* 2

Now suppose for some 𝓁 *≥* 2, Λ_𝓁_ = {Γ (*𝒢*; *d, h*) : *d, h}*. We aim to show that Λ _𝓁 +1_ = {Γ_+1_(*𝒢*; *d, h*) : *d, h}*. Consider an arbitrary element Γ _𝓁 +1_(*d*^†^, *h*^†^) ∈ {Γ_+1_(*𝒢*; *d, h*) : *d, h}*.

Then

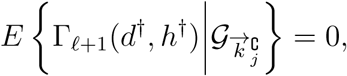

for *j* = 1, …, 𝓁 + 1 by (S3), and therefore Γ_𝓁 +1_(*d*^†^, *h*^†^) ∈ Λ_𝓁 +1_. Conversely, let *ω* ∈ Λ_𝓁 +1_, which implies that

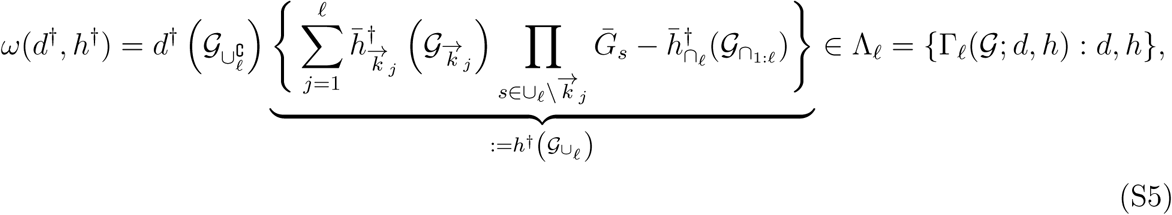

for some index functions (*d*^†^, *h*^†^), and also

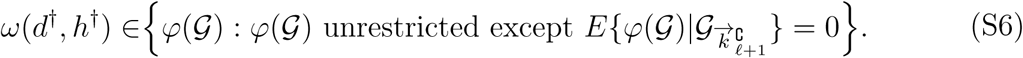

We consider the following two cases:

**Case 1**: {*∪*_𝓁: +1_*\∪*_1: 𝓁_ } is a singleton

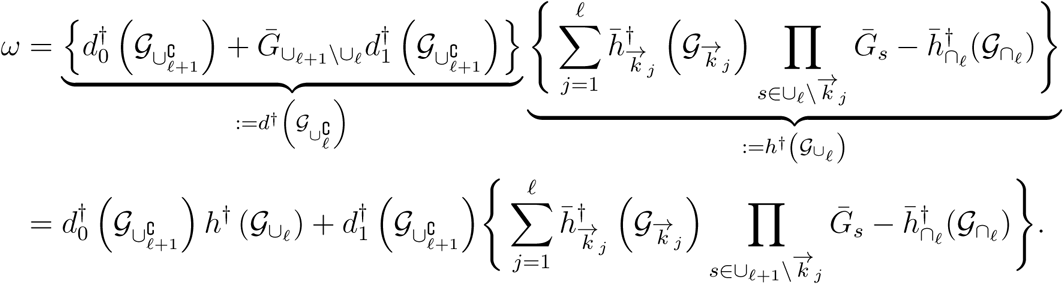

Following the proof of Proposition 3, we have

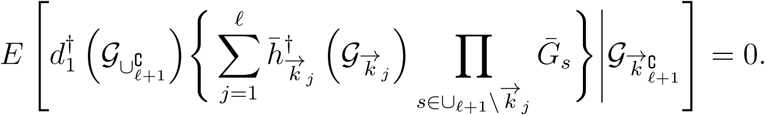

Then (S6) has the nontrivial implication that

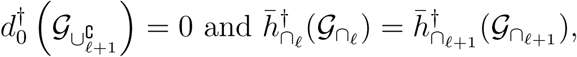

almost surely, so that *ω* ∈ {Γ_𝓁 +1_(𝒢; *d, h*) : *d, h}*.

**Case 2:** *∪*_𝓁 +1_ = *∪*_𝓁_

In this case,

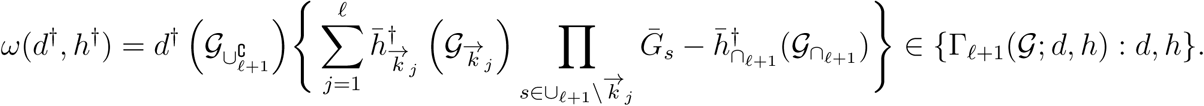

We may conclude that Λ_𝓁 +1_ = {Γ_𝓁 +1_(𝒢; *d, h*) : *d, h*}, and (S4) holds by mathematical induction. In particular Λ_*K*_ = {Γ_*K*_(𝒢; *d, h*) : *d, h*} where

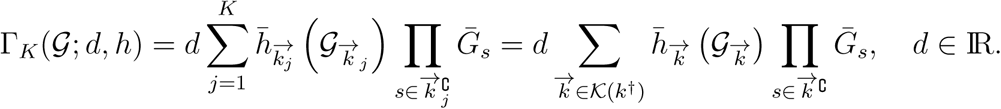

Hence,

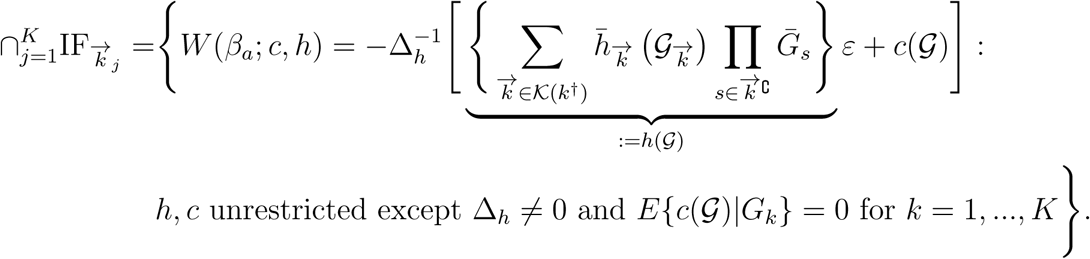

## Semiparametric Efficiency Bound of *β*_*a*_ in the Union Model ℳ_*union,k*_†

For any *c*(𝒢) ≠ 0 almost surely, the difference of the asymptotic variances of the estimators indexed by *W* (*β*_*a*_; *c, h*) and *W* (*β*_*a*_; *c* = 0, *h*) respectively for fixed *h*(𝒢) is

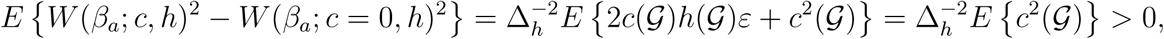

where the last equality follows from

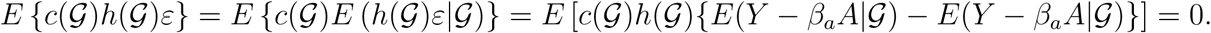

Therefore the efficient influence function of *β*_*a*_ lies in the set

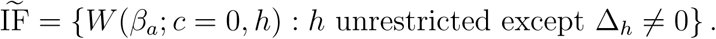

We derive the efficient element in 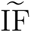 by considering saturated models for the functions 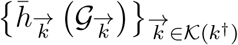 in *h*(𝒢). Then using a symmetry argument, the index function *h*(𝒢) = *θ*^*T*^ **H**(*G*) is a linear combination with parameter vector 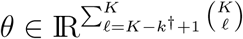 and a vector of basis functions 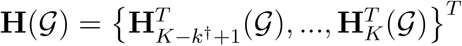 where for each 𝓁 = *K − K*^†^ + 1,…,K,

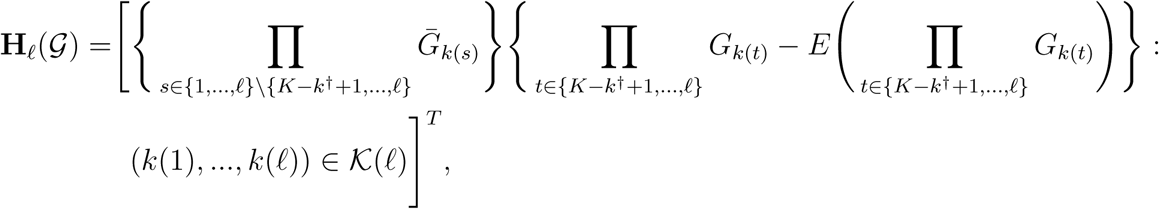

is a 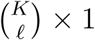 vector of *ℓ*-way interactions. Therefore we have the equivalent representation

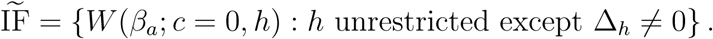

By Theorem 5.3 of Newey and McFadden (1994), the optimal linear combination in terms of asymptotic variance is indexed by 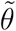 which satisfies the generalized information equality

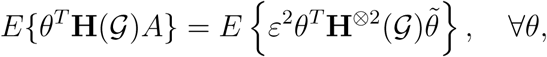

where *Q*^*⊗*2^ := *QQ*^*T*^. Equivalently,

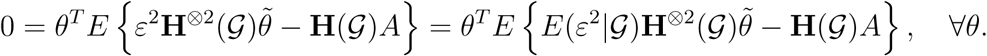

Taking 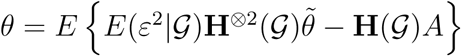 yields

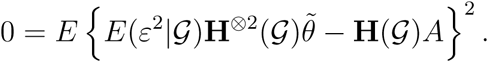

Hence, 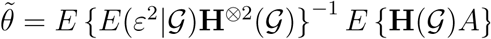 and

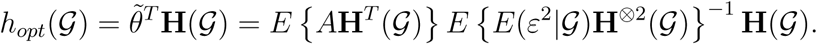

The semiparametric efficiency bound of *β*_*a*_ in ℳ_*union,k*_† is therefore given by

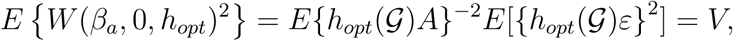

where *V* = [*E* {*A***H**^T^ (𝒢) *E* {*E*(ε^2^|𝒢) **H** ^⊗2^ (𝒢) }^− 1^ *E* {**H** (𝒢)*A* }]^− 1^ In particular when 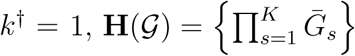 consists of only one basis function and

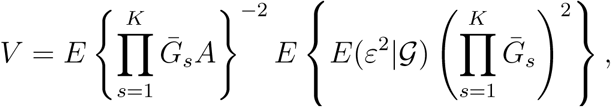

a result given in Theorem 1.

## Influence Function of the MR^2^ Estimator

Let **Z**(𝒢) denote a user-specified vector of mean zero basis functions, evaluated at {*E*(*G*_*j*_)}_*j*=1,…,*K*_. For the example in the manuscript,

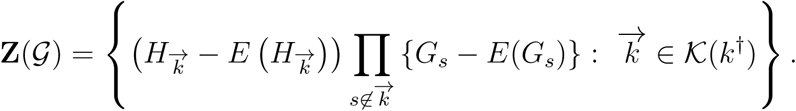

In addition, let 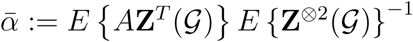 denote the population exposure regression coefficients. It follows from the results of Robins (2000) and Okui et al. (2012) that the two-stage least squares estimator 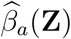 has the asymptotic expansion

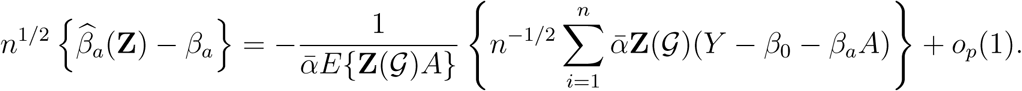

Let 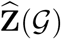 denote the same user-specified vector of basis functions, but evaluated at the (non-parametrically) estimated means {*Ê* (*G*_*j*_)}_*j*=1,…,*K*_. Following Newey (1994), the adjustment term to the above influence function due to nonparametric estimation of the means in the semiparametric union model ℳ_*union,k*_† is 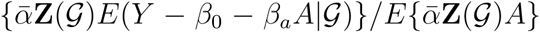, and therefore

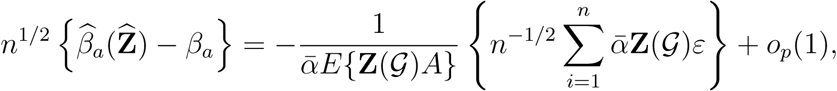

with influence function *–*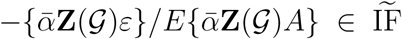. By Slutsky’s Theorem and the Central Limit Theorem,

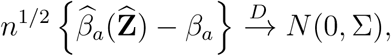

as *n → ∞*, where 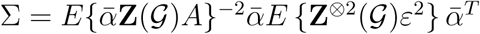. The standard sandwich estimator

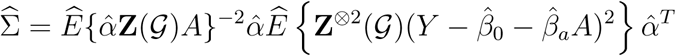

of Σ which ignores first stage estimation of {*E*(*G*_*j*_)}_*j*=1,…,*K*_, is readily available from existing off-the-shelf software. Because Cov {**Z**(𝒢)(*Y −β*_0_ *−β*_*a*_*A*)*}−E* [Cov {**Z**(𝒢)(*Y −β*_0_ *−β*_*a*_*A*)| 𝒢}] is positive semidefinite, 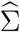 is a conservative estimator of the asymptoptic variance. An appealing feature of the MR^2^ estimator under an additional homoskedasticity assumption *E*(*ε*^2^| 𝒢) = *E*(*ε*^2^) := *σ*^2^ is that it is the efficient one among the class of estimators that uses linear combinations of the basis functions in **Z**(𝒢) (see Section 5.2.3 of Wooldridge (2010)). In this case the asymptotic variance formula simplifies to

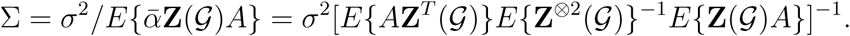

In particular, if the analyst uses the full suite of basis functions **H**(𝒢), then Σ = *V* and the MR^2^ estimator attains the semiparametric efficiency bound asymptotically.

## Proof of Proposition 3

Let 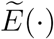 denote the expectation with respect to the probability mass function 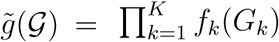. Because 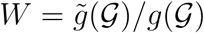 is a function of 𝒢 only, it follows from the proof of Proposition 2 that

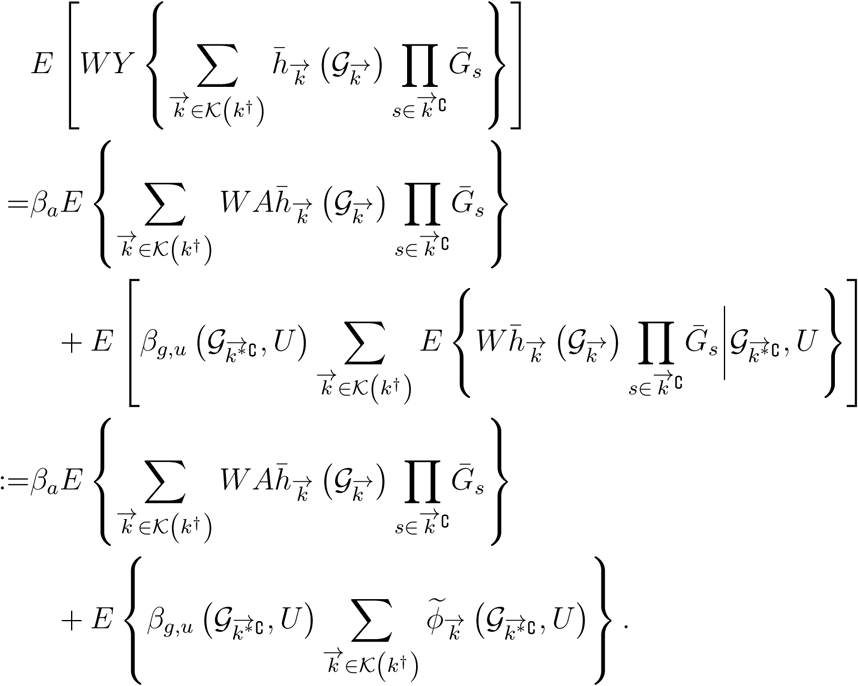

Consider each term 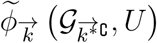 If 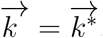, then

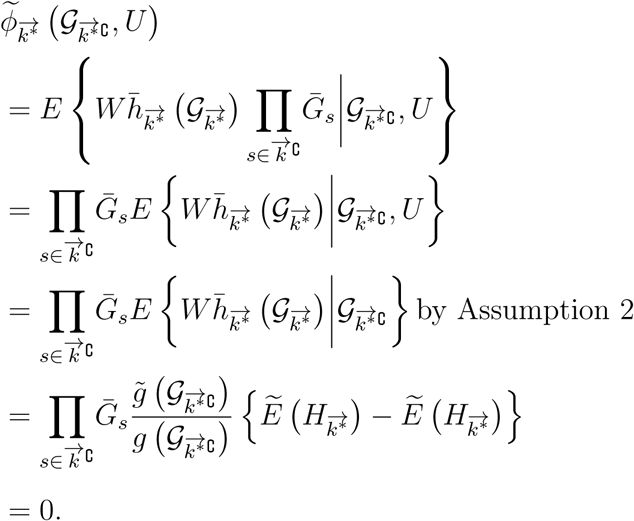

Otherwise 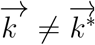, and

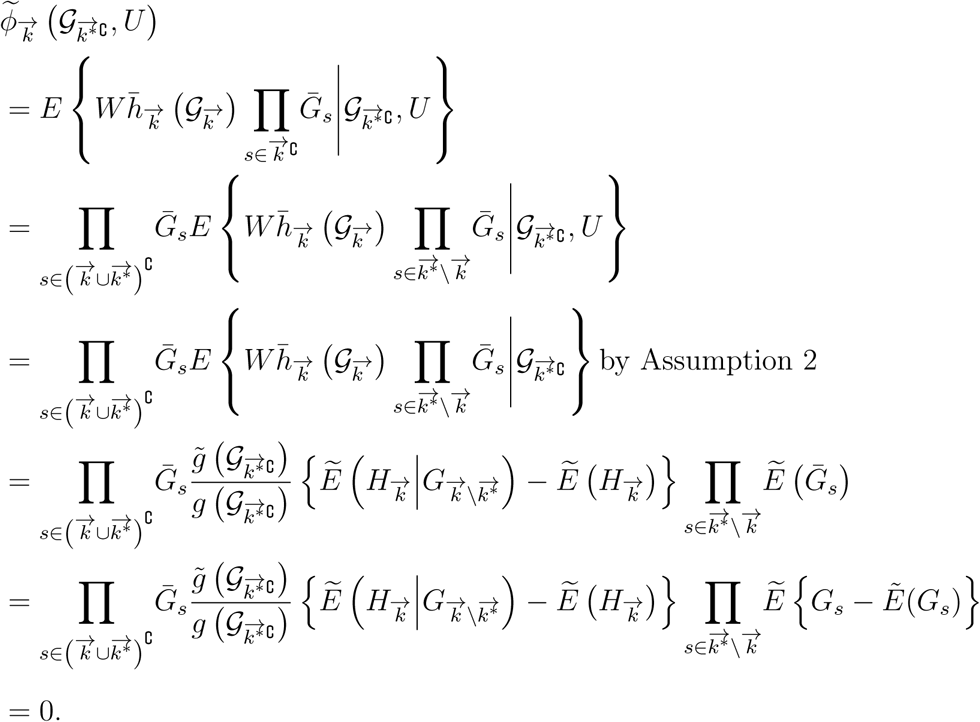

Therefore 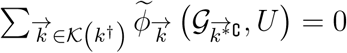 almost surely,and

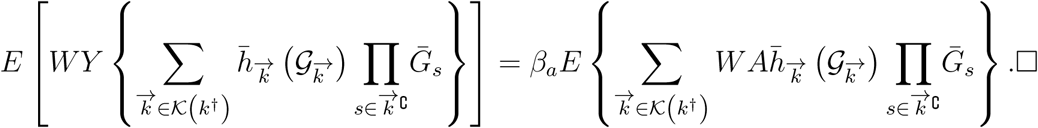

## Additional Simulation Results

**Table S1:** Comparison of methods with continuous exposure generated under the identity link (*C* = 1.0). The two rows of results for each estimator correspond to sample sizes of *n* = 10000 and *n* = 50000 respectively.

|  | MR<sup>2</sup> | Oracle 2SLS | Naive 2SLS | Post-Lasso | TSHT |
| --- | --- | --- | --- | --- | --- |
| 50% rule holds |  |  |  |  |  |
| \|Bias\| | .004 | .000 | .008 | .000 | .000 |
|  | .001 | .000 | .009 | .000 | .000 |
| $\sqrt{\text{Var}}$ | .012 | .001 | .001 | .002 | .002 |
|  | .006 | .001 | .001 | .001 | .001 |
| $\sqrt{\text{EVar}}$ | .011 | .001 | .001 | .002 | .001 |
|  | .006 | .001 | .001 | .001 | .001 |
| Cov95 | .931 | .951 | .000 | .939 | .950 |
|  | .941 | .945 | .001 | .947 | .947 |
| 50% rule violated but plurality rule holds |  |  |  |  |  |
| \|Bias\| | .018 | .000 | .013 | .009 | .005 |
|  | .010 | .000 | .013 | .007 | .000 |
| $\sqrt{\text{Var}}$ | .035 | .002 | .001 | .005 | .005 |
|  | .026 | .001 | .001 | .004 | .002 |
| $\sqrt{\text{EVar}}$ | .041 | .002 | .001 | .005 | .005 |
|  | .028 | .001 | .001 | .001 | .001 |
| Cov95 | .937 | .937 | .000 | .119 | .425 |
|  | .936 | .951 | .000 | .000 | .944 |
| Both 50% and plurality rules violated |  |  |  |  |  |
| \|Bias\| | .019 | .000 | .013 | .021 | .021 |
|  | .010 | .000 | .013 | .021 | .021 |
| $\sqrt{\text{Var}}$ | .035 | .002 | .001 | .002 | .002 |
|  | .026 | .001 | .001 | .001 | .001 |
| $\sqrt{\text{EVar}}$ | .041 | .002 | .001 | .001 | .002 |
|  | .028 | .001 | .001 | .001 | .001 |
| Cov95 | .936 | .940 | .000 | .000 | .000 |
|  | .936 | .950 | .000 | .000 | .000 |

Note: |Bias| and $\sqrt{\text{Var}}$ are the Monte Carlo absolute bias and standard deviation of the point estimates, $\sqrt{\text{EVar}}$ is the square root of the mean of the variance estimates and Cov95 is the coverage proportion of the 95% confidence intervals, based on 1000 repeated simulations. Zeros denote values smaller than .0005.

**Table S2:**
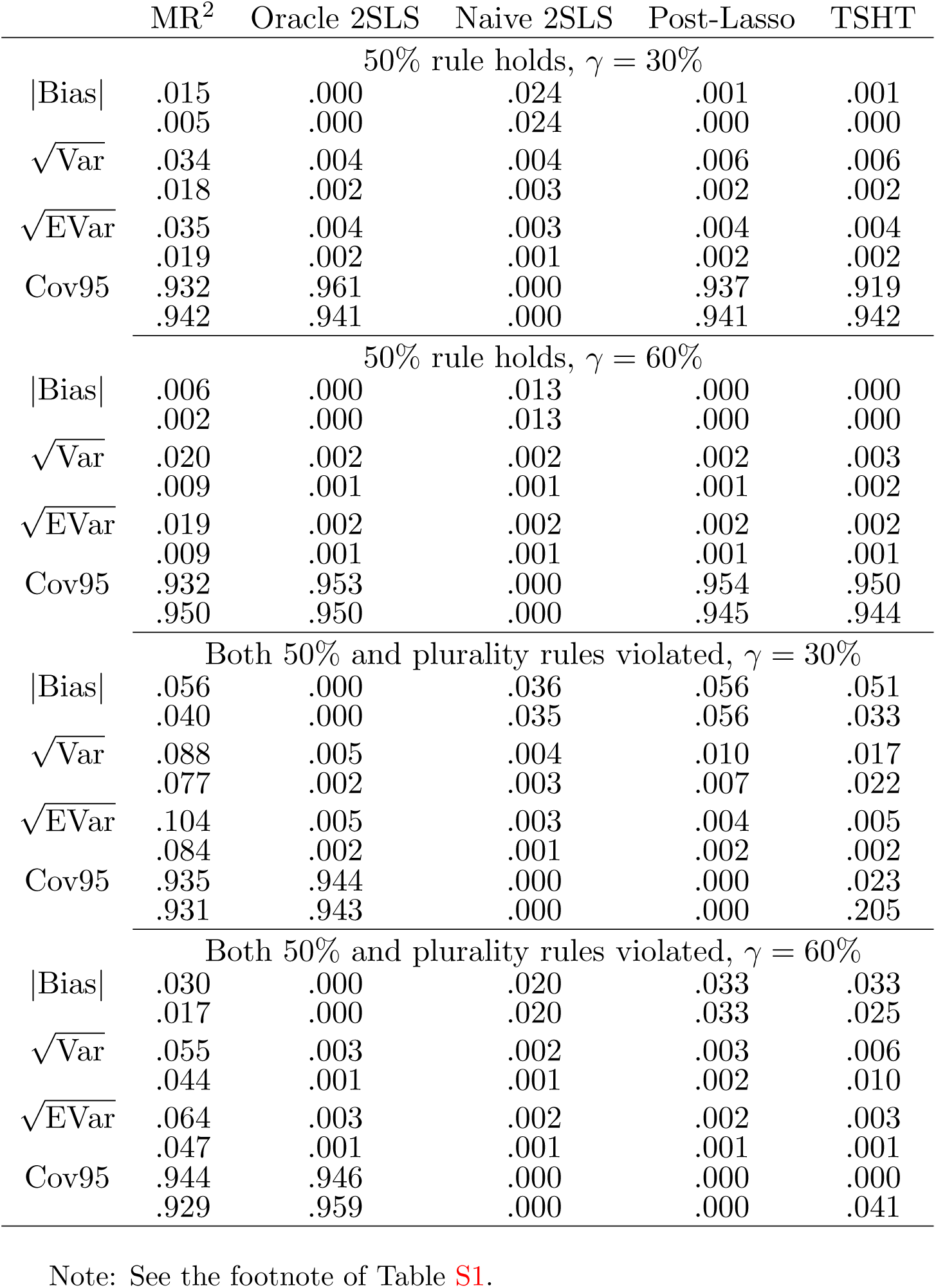
Comparison of methods with continuous exposure generated under the identity link (*C* = 1.0), and varying degree of interactive effects. The two rows of results for each estimator correspond to sample sizes of *n* = 10000 and *n* = 50000 respectively.

## UK Biobank Data Analysis

The following table provides detailed information about the 10 SNPs used in the main text for estimating the average causal effects of BMI on SBP and DBP respectively. These 10 SNPs are independent from each other.

**Table S3:** Information about the top 10 SNPs used in the UK Biobank data analysis in the main text. CHR stands for chromosome, BP stands for the base-pair position in the chromosome, EAF stands for the allele frequency of allele 1 (effect allele), Var-ex stands for the variance explained by the SNP, F-stat stands for the F-statistics in the first -stage regression.

| SNP Name | CHR | BP | Gene | Allele 1/2 | F-stat | Effect | SE | EAF | Var-ex | P-value |
| --- | --- | --- | --- | --- | --- | --- | --- | --- | --- | --- |
| rs1558902 | 16 | 52361075 | FTO | A/T | 781.20 | 0.082 | 0.003 | 0.415 | 0.33% | 7.51E-153 |
| rs6567160 | 18 | 55980115 | MC4R | C/T | 298.67 | 0.056 | 0.004 | 0.236 | 0.11% | 3.93E-53 |
| rs13021737 | 2 | 622348 | TMEM18 | G/A | 229.98 | 0.06 | 0.004 | 0.828 | 0.10% | 1.11E-50 |
| rs10938397 | 4 | 44877284 | GNPDA2 | G/A | 221.47 | 0.04 | 0.003 | 0.434 | 0.08% | 3.21E-38 |
| rs543874 | 1 | 176156103 | SEC16B | G/A | 189.54 | 0.048 | 0.004 | 0.193 | 0.07% | 2.62E-35 |
| rs2207139 | 6 | 50953449 | TFAP2B | G/A | 155.24 | 0.045 | 0.004 | 0.177 | 0.06% | 4.13E-29 |
| rs11030104 | 11 | 27641093 | BDNF | A/G | 147.65 | 0.041 | 0.004 | 0.792 | 0.06% | 5.56E-28 |
| rs3101336 | 1 | 72523773 | NEGR1 | C/T | 129.01 | 0.033 | 0.003 | 0.613 | 0.05% | 2.66E-26 |
| rs7138803 | 12 | 48533735 | BCDIN3D | A/G | 126.37 | 0.032 | 0.003 | 0.384 | 0.05% | 8.15E-24 |
| rs10182181 | 2 | 25003800 | ADCY3 | G/A | 120.26 | 0.031 | 0.003 | 0.462 | 0.05% | 8.78E-24 |

**Figure S1:**
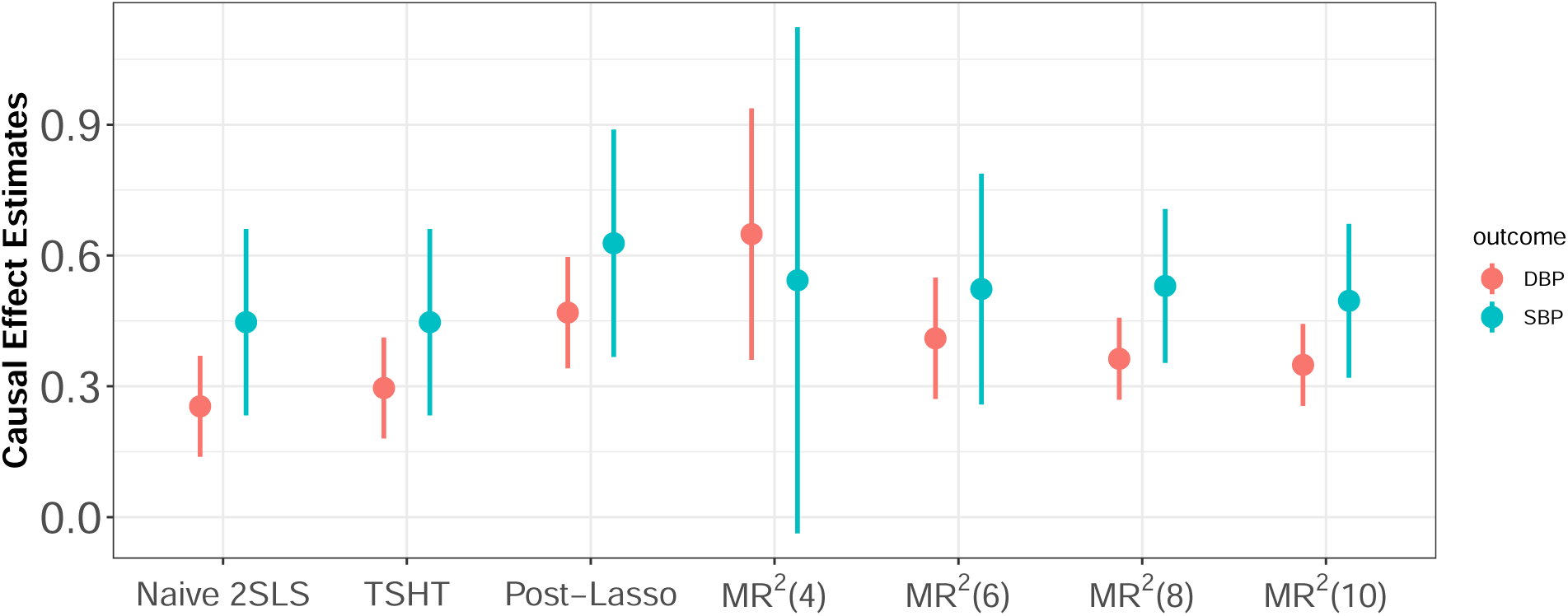
Point estimates (solid dots) of the causal effects of BMI on SBP and DBP with corresponding 95% confidence intervals (vertical bars) respectively.

## Footnotes

1 The sets 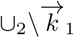 and 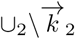 are singletons due to the enumeration scheme.

